# Emergence and phenotypic characterization of C.1.2, a globally detected lineage that rapidly accumulated mutations of concern

**DOI:** 10.1101/2021.08.20.21262342

**Authors:** Cathrine Scheepers, Josie Everatt, Daniel G. Amoako, Houriiyah Tegally, Constantinos Kurt Wibmer, Anele Mnguni, Arshad Ismail, Boitshoko Mahlangu, Bronwen E. Lambson, Simone I. Richardson, Darren P. Martin, Eduan Wilkinson, James Emmanuel San, Jennifer Giandhari, Nelia Manamela, Noxolo Ntuli, Prudence Kgagudi, Sandile Cele, Sureshnee Pillay, Thabo Mohale, Upasana Ramphal, Yeshnee Naidoo, Zamantungwa T. Khumalo, Gaurav Kwatra, Glenda Gray, Linda-Gail Bekker, Shabir A. Madhi, Vicky Baillie, Wesley C. Van Voorhis, NGS-SA, Florette K. Treurnicht, Marietjie Venter, Koleka Mlisana, Nicole Wolter, Alex Sigal, Carolyn Williamson, Nei-yuan Hsiao, Nokukhanya Msomi, Tongai Maponga, Wolfgang Preiser, Zinhle Makatini, Richard Lessells, Penny L. Moore, Tulio de Oliveira, Anne von Gottberg, Jinal N. Bhiman

## Abstract

Global genomic surveillance of SARS-CoV-2 has identified variants associated with increased transmissibility, neutralization resistance and disease severity. Here we report the emergence of the PANGO lineage C.1.2, detected at low prevalence in South Africa and eleven other countries. The emergence of C.1.2, associated with a high substitution rate, includes changes within the spike protein that have been associated with increased transmissibility or reduced neutralization sensitivity in SARS-CoV-2 VOC/VOIs. Like Beta and Delta, C.1.2 shows significantly reduced neutralization sensitivity to plasma from vaccinees and individuals infected with the ancestral D614G virus. In contrast, convalescent donors infected with either Beta or Delta showed high plasma neutralization against C.1.2. These functional data suggest that vaccine efficacy against C.1.2 will be equivalent to Beta and Delta, and that prior infection with either Beta or Delta will likely offer protection against C.1.2.

---

More than a year into the COVID-19 pandemic, SARS-CoV-2 remains a global public health concern with ongoing waves of infection resulting in the selection of SARS-CoV-2 variants with novel constellations of mutations within the viral genome^1–3^. Some variants accumulate mutations within the spike region that result in increased transmissibility and/or immune evasion, making them of increased public health importance^1–3^. Depending on their clinical and epidemiological profiles, these are either designated as variants of interest (VOI) or variants of concern (VOC) (www.who.int), and ongoing genomic surveillance is essential for early detection of such variants. At the time of writing there were four VOCs (Alpha, Beta, Gamma and Delta) and five VOIs (Eta, Iota, Kappa, Lambda and Mu) in circulation globally (www.who.int, accessed September 21, 2021). Alpha, Beta and Delta have had the most impact in terms of transmission and immune evasion, with Delta rapidly displacing other variants to dominate globally and in South Africa^4^.

Ongoing genomic surveillance by the Network for Genomic Surveillance in South Africa (NGS-SA)^5^ detected an increase in sequences assigned to C.1 during the country’s third wave of SARS-CoV-2 infections from March 2021. This was unexpected since C.1, first identified in South Africa^6,7^, was last detected in January 2021. Upon comparison of the mutational profiles between the new and older C.1 sequences (which only contain the D614G spike mutation), it was clear that the new sequences had mutated substantially. C.1 had minimal spread globally, but was detected in Mozambique where it was found to have accumulated additional mutations, resulting in the PANGO lineage C.1.1^7^. The new C.1-like sequences discovered from March 2021 were, however, very distinct from C.1.1, and were subsequently assigned to the PANGO lineage C.1.2.

As of data deposited on September 10, 2021 we identified 166 sequences that match the C.1.2 lineage (available on GISAID (www.gisaid.org), the global reference database for SARS-CoV-2 viral genomes, and listed in **Supplementary Tables 1** and **2**). While these 166 sequences make up the C.1.2 dataset used in this analysis, at the time of submission (September 22, 2021) a further 11 C.1.2 sequences had been deposited in GISAID, including detection in the United States of America. The majority of the 166 sequences (n=146, including 10 from vaccine breakthrough cases (**Supplementary Table 1**)) are from South Africa. The remaining sequences were detected in Botswana, China, the Democratic Republic of the Congo (DRC), Eswatini, England, Mauritius, New Zealand, Portugal, Switzerland and Zimbabwe, with at least 11/20 cases having a travel history from South Africa (**Supplementary Fig. 1a** and **Supplementary Table 2**). Provincial detection of C.1.2 mirrored the depth of sequencing across South Africa (**Supplementary Fig. 1b, c** and **d**), suggesting that the observed numbers may be an underrepresentation of the spread and frequency of this variant within South Africa and globally. We observed increased detection rates of C.1.2 in May, June and July (**Supplementary Fig. 1e**). Although the C.1.2 detection rate decreased in August, very few sequences have been obtained for the end of the month based on data submitted to GISAID as of September 21, 2021 (**Supplementary Fig. 1d**). Nevertheless, these rates are similar to the increases seen at the start of Beta and Delta detection in South Africa, though notably a sudden large increase in C.1.2 has not yet occurred (**Supplementary Fig. 1e**). Spatiotemporal phylogeographic analysis predicts the root of the C.1.2 lineage around mid-July 2020 (95% highest posterior density ranging from the May to October 2020) in Gauteng or surrounding provinces (**Supplementary Fig. 1f**). Onward spread from Gauteng to neighbouring provinces is inferred to have occurred in early 2021, while introduction to more distant provinces occurred from the end of May 2021.

C.1.2 is highly mutated, even when compared to circulating VOI/VOCs (**Fig. 1a**). The emergence of the Alpha, Beta, Delta and Gamma variants were associated with short periods of notably increased evolution compared to the overall SARS-CoV-2 evolutionary rate^8^. The same is observed for C.1.2, which has an approximate evolutionary rate of 3.04×10^−3^ nucleotide changes/site/year (**Fig. 1b**). The short periods of increased evolution associated with the emergence of this and other new lineages are indicative of the accumulation of mutations having occurred during discrete epidemiological events, such as virus-host co-evolution within individuals with prolonged viral infections^9,10^, or genetic recombination between two distantly related viral variants co-infecting the same individual. Although C.1.2 shares multiple mutations with the Alpha, Beta, and Delta variants, we were unable to find evidence for recombination events between these VOCs or C.1 within C.1.2 viruses (**Supplementary Fig. 2c**). Both the absence of evidence of recombination, and the phylogenetic placement of C.1.2 relative to the other VOCs, support the hypothesis that the mutations in C.1.2 likely occurred as a result of convergent evolution within the context of prolonged infection within an individual^9,10^.

**Figure 1:**
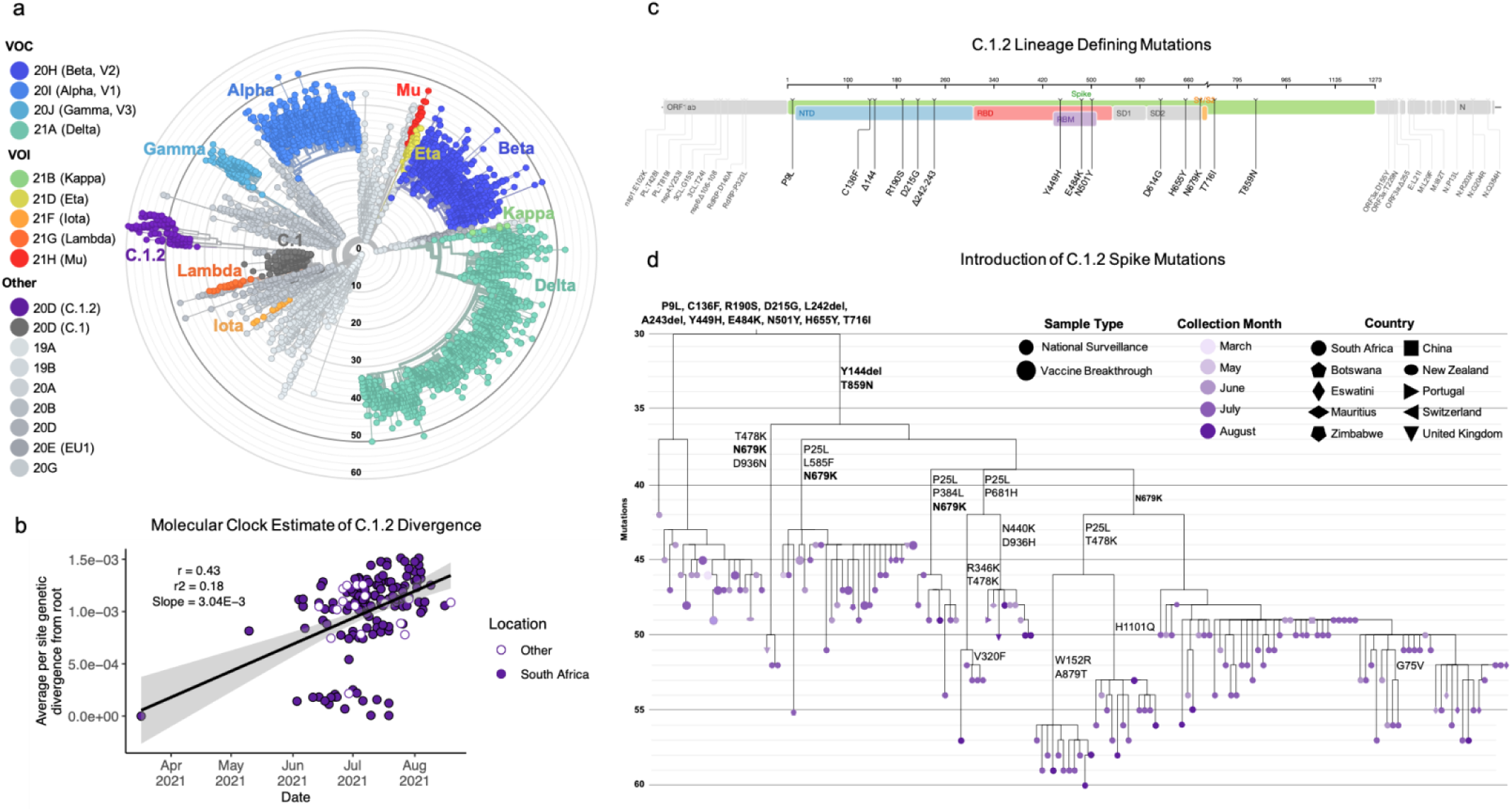
C.1.2 a highly mutated SARS-CoV-2 variant. **a**, Phylogenetic tree of 6,192 global sequences (1,991 sequences from South Africa), including Variants of Concern (VOC), Variants of Interest (VOI), and the C.1.2 lineage, colored by Nextstrain clade (shown in the key) and scaled by divergence (number of mutations). The C.1.2 lineage (purple) forms a distinct, highly mutated cluster within clade 20D. **b**, Regression of root-to-tip genetic distances against sampling dates for sequences belonging to lineage C.1.2 sampled either in South Africa (solid purple) or in other countries (white) indicating that the C.1.2 sequences evolved in a clock-like manner (correlation coefficient = 0.43, R2 = 0.18). The regression gradient is an estimate of the rate of sequence evolution, which is 3.04×10-3 nucleotide substitutions/site/year. **c**, Full genome representation of C.1.2 showing lineage defining mutations (seen in ≥50% of C.1.2 assigned sequences), with those in the spike (green) colored according to functional regions, including the N-terminal domain (NTD, blue), receptor binding domain (RBD, red), receptor binding motif (RBM, purple), subdomain 1 and 2 (SD1 or SD2, grey) and the cleavage site (S1/S2, yellow). Figure generated by covdb.stanford.edu. **d**, A phylogenetic tree highlighting the introduction of spike mutations in the different sub-clades of the C.1.2 lineage. The tree is coloured by month of collection, with tip symbols indicating country of collection, as indicated in the key (Figure generated from a Nextstrain build of global C.1.2 sequences with ≥90% coverage). Mutations in bold represent those observed in ≥50% of the C.1.2 sequences.

C.1.2 has accumulated additional mutations within the ORF1ab, spike, ORF3a, ORF9b, E, M and N genes compared to its C.1 ancestor (**Fig. 1c**). C.1.2 has, on average, 30 amino acid substitutions across its entire genome, similar to the frequency observed in Delta (average: 28) (**Supplementary Fig. 2a**). On average, 13 of these amino acid substitutions were observed within the spike protein, notably higher than Delta (average: 7) but similar to Gamma (average: 12) (**Supplementary Fig. 2a**). The spike mutations observed in C.1.2 include five within the N-terminal domain (NTD: C136F, Y144del, R190S, D215G and 242-243del or 243-244del (either deletion results in the same amino acid sequence), three within the receptor binding motif (RBM: Y449H, E484K and N501Y) and three adjacent to the furin cleavage site (H655Y, N679K and T716I) (**Fig. 1c**). Though these mutations, along with P9L and T859N, occur in the majority of C.1.2 viruses, there is additional variation within the spike region of this lineage (**Supplementary Fig. 2b and c**). Additional mutations include P25L (in ∼43% of viruses) and W152R (in ∼7%) in the NTD, T478K (∼17%) in the RBM, L585F (∼17%) in S1, P681H (∼8%) adjacent to the furin cleavage site, A879T (∼7%), D936H (∼5%), and H1101Q (∼8%) in S2, with additional mutations detected in less than 5% of viruses (**Supplementary Fig. 2b**). Though the prevalence of some of these mutations may be underestimated due to lack of sequencing coverage in parts of the S gene (**Supplementary Fig. 2c**), their presence, particularly in the most recent samples, suggests ongoing intra-lineage evolution. The majority of these mutations (P9L, C136F, R190S, D215G, L242del, A243del, Y449H, E484K, N501Y, H655Y, and T716I), however, appeared simultaneously, further supporting a single, prolonged infection giving rise to this lineage (**Fig. 1d**).

Of the 28 spike mutations identified in C.1.2, 14 (50%) have been previously identified in other VOIs and VOCs, including mutations associated with increased transmissibility and/or neutralization escape (**Fig. 2a**). These mutations include D614G, common to most circulating SARS-CoV-2 variants and associated with increased viral fitness^11^, and E484K and N501Y in the RBD, which are shared with multiple VOCs and VOIs and associated with reduced antibody responses^12,13^. C.1.2 contains another two RBD mutations not seen in other VOIs or VOCs: N440K and Y449H, which co-localize on the same outer face of the RBD (**Fig. 2b**). While these two mutations are not characteristic of current VOCs/VOIs, they have been associated with escape from certain class 3 neutralizing antibodies^14^. The Y144del and 242-244del cause shifts in the immunodominant N3 and N5 loops of the NTD (blue, **Fig. 2b**) in the Alpha and Beta variants, respectively, altering antigenicity of the region^12^. Recurrent NTD deletions have also previously been associated with prolonged SARS-CoV-2 infection^15^. Furthermore, the C136F mutation abolishes a disulphide bond within the N1 loop of NTD, and in combination with P25L likely contributes to immune escape by conformationally liberating the entire N-terminus of the NTD. The P9L within the signal peptide is predicted to increase translation. Mutations close to the furin cleavage site, including H655Y and P681R/H, have been shown to increase S1/S2 cleavage efficiency, and have also been observed in other VOCs, such as Alpha, Delta, Gamma and Kappa^16–18^ (S1/S2 region in **Fig. 2b**). In the C.1.2 lineage, N679K and P681H are mutually exclusive (with N679K predominating, **Fig. 1d**), which suggests that they may perform a similar role by increasing the local, relative positive charge of the furin cleavage site to potentially improve the furin cleavage efficiency. Similar mutations have been seen within Gamma^19^. Finally, T859N has been detected in multiple non-VOC/VOI lineages and is predicted to affect spike stability in a similar way as the D164G substitution. The identification of convergent evolution between C.1.2 and the presently designated VOIs and VOCs suggests that this lineage may also share concerning phenotypic properties with these VOIs and VOCs.

**Figure 2:**
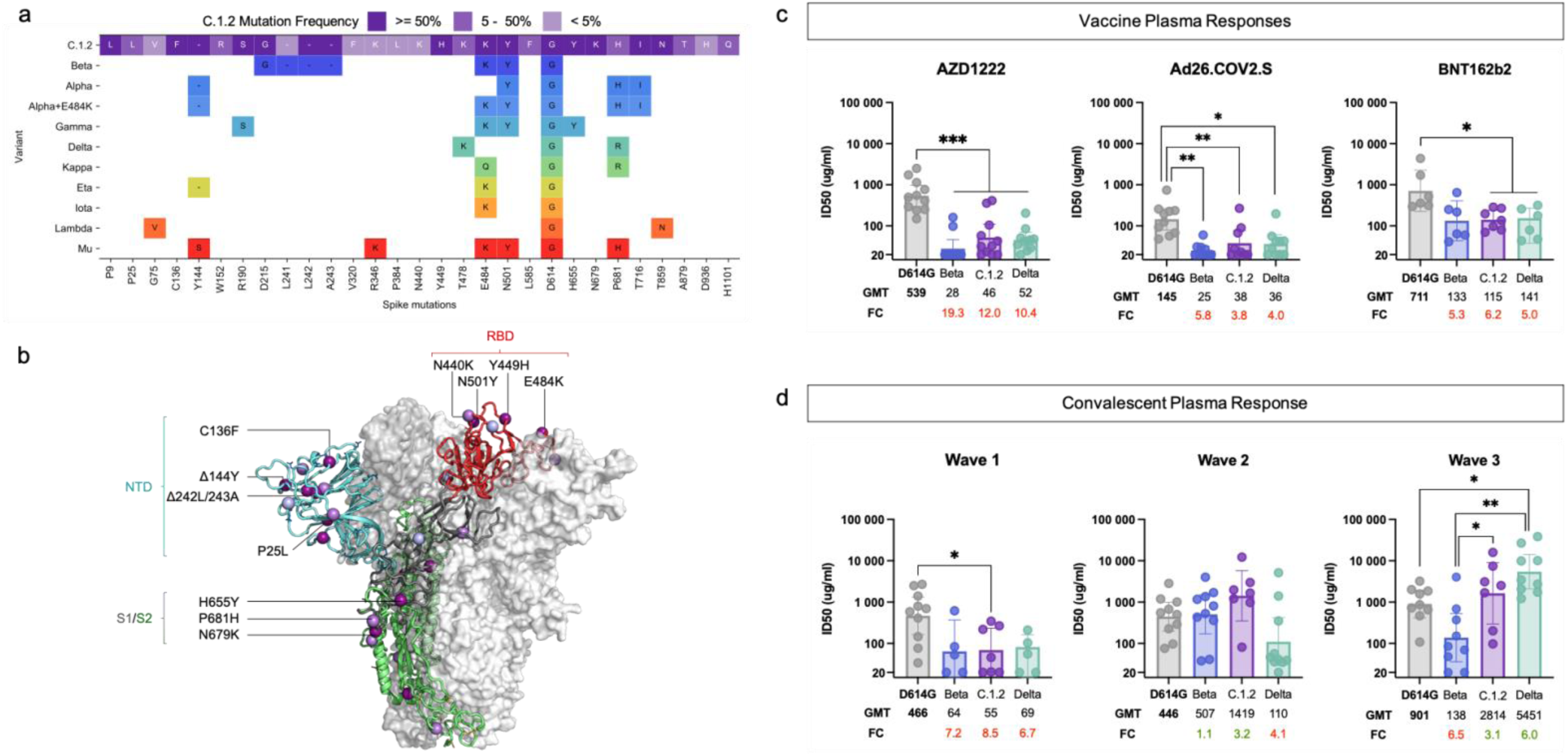
Shared spike mutation results in similar immune responses. **a**, Visualization of C.1.2 spike mutations, highlighting those shared with VOCs and VOIs (colored by the Nextstrain clade). All C.1.2 mutations are shown, and coloured according to prevalence within the C.1.2 sequences (shown in key). For VOCs and VOIs only mutations present in at least 50% of sequences are shown (as determined by frequency information at outbreak.info, accessed September 13, 2021). **b**, Schematic showing C.1.2 mutations on the RBD-down conformation of SARS-CoV-2 spike, with domains of a single protomer shown in cartoon view and colored cyan (N-terminal domain, NTD), red (C-terminal domain/receptor binding domain, CTD/RBD), grey (subdomain 1 and 2, SD1 and SD2), and green (S2). The adjacent protomers are shown in translucent surface view and colored shades of grey. Lineage-defining mutations (found in >50% of sequences) are coloured dark purple, with additional mutations (present in <50% of sequences) coloured light purple. Key mutations known/predicted to influence neutralization sensitivity (C136F and P25L, Δ144Y, Δ242L/243A, and E484K), or furin cleavage (H655Y and N679K) are indicated. Image was created using the PyMOL molecular graphic program. **c** and **d**, Neutralization activity of plasma samples taken from donors previously vaccinated with either AZD1222, Ad26.COV.2.S or BNT162b2 (panel **c**) and patients previously infected during the first, second or third waves in South Africa (panel **d**) against the wild-type (D614G), Beta, Delta and C.1.2 variants. Bar graphs represent the geometric mean titer (GMT) for each group with the error bars representing the 95% confidence intervals, dots represent individual sample titers. Statistical significance based on the Wilcoxon test are shown above graphs. P-values are denoted with “*” symbols: * p<0.05, ** p<0.01 and *** p<0.001. GMT and fold-change (FC) differences relative to D614G (shown in bold) are given below the graph, with red representing decreased titre and green representing increased titre.

Plasma neutralizing activity from donors who received the ChAdOx1 nCoV-19 (AZD1222), Jansen/Johnson and Johnson (Ad26.COV2.S) or Pfizer/BioNTech (BNT162b2) vaccines, showed 3 to 12-fold reduction in antibody titres for C.1.2 compared to the original D614G variant (containing only the D614G spike mutation). Despite this, no statistically significant difference in the reduction in titers against Beta (5 to 19-fold) or Delta (4 to 10-fold) was observed (**Fig. 2c** and **Supplementary Fig. 3a**). Similarly, convalescent plasma from donors infected with the D614G variant that dominated the first wave of infections in South Africa^6^ showed reduced responses to Beta (geometric mean titre, GMT of 64), C.1.2 (GMT: 55) and Delta (GMT: 69) (**Fig. 2d**, left panel and **Supplementary Fig. 3b**). Convalescent plasma from donors infected during the second wave of infections in South Africa (dominated by Beta), showed reduced sensitivity against Delta (GMT: 110) but high neutralizing activity against C.1.2 (GMT: 1419). This may be attributed to the shared E484K and N501Y RBD mutations in Beta and C.1.2 variants, which are absent in Delta (**Fig. 2d**, middle panel and **Supplementary Fig. 3a**, middle panel). In donors infected during South Africa’s third wave of Delta-dominated infections^4^, neutralizing activity was reduced against D614G and Beta, but relatively high titres were observed against C.1.2 (GMT: 2814) (**Fig. 2d**, right panel and **Supplementary Fig. 3a**, right panel). High neutralization titres against D614G, C.1.2 and Delta are likely a result of higher viral loads associated with Delta infections^20^, while reduced titres against Beta may be a result of the K417N substitution, which escapes a predominant antibody class^12^, and is not present in Delta or C.1.2. Antibody dependent cellular cytotoxicity (ADCC) in both vaccinees and wave one convalescent donors was significantly reduced against C.1.2 relative to D614G. (**Fig. 2c** and **Supplementary Fig. 3b**). In contrast, as with neutralization, ADCC responses against C.1.2 in convalescent plasma from individuals infected in waves two and three, were largely preserved (**Supplementary Fig. 3b**).

Overall these results show that, as with Beta and Delta, vaccine-induced antibody responses show reduced activity against the C.1.2 variant. However, prior infection with either Beta or Delta will likely confer some protection against C.1.2. Cross-reactivity between C.1.2 and Beta/Delta could be a result of shared mutations between these viruses. Though C.1.2 is present in South Africa and globally, we have not yet seen exponential expansion of this lineage as was observed prior to local Beta and Delta dominance^4^. This could be a result of recent increased population immune protection through cross-reactive antibodies induced by prior Beta or Delta infections against C.1.2.

## Data Availability

All SARS-CoV-2 assemblies used in this analysis are deposited in GISAID (https://www.gisaid.org/)20,21 and the GISAID accessions are provided in Supplementary Tables 1 and 2. The Nextstrain build of C.1.2 and global sequences will be made available at https://nextstrain.org/groups/ngs-sa.

https://www.gisaid.org/

https://nextstrain.org/groups/ngs-sa.

## Acknowledgements

We acknowledge additional NGS-SA members: Adriano Mendes, Allison Glass, Amy Strydom, Arash Iranzadeh, Bulelani Manene, Cheryl Cohen, Deelan Doolabh, Derek Tshiabuila, Diana Hardie, Dominique Goadhals, Gert van Zyl, Innocent Madau, Kamela Mahlakwane, Kathleen Subramoney, Kruger Marais, Linda de Gouveia, Lynn Tyers, Michaela Davids, Noluthando Duma, Rageema Joseph, Yajna Ramphal, Upasana Ramphal, Sibongile Walaza, Simnikiwe Mayaphi, Stephen Korsman, Susan Engelbrecht, Tania Stander, Terry Marshall, Zinhle Makatini. We thank Clare Cutland and Anthonet Koen from the South African Medical Research Council Vaccines and Infectious Diseases Analytics Research Unit for providing the AZD1222 samples. We acknowledge Sasha W. Tiles for the provision of Pfizer plasma samples for the United World Antivirus Research Network (UWARN). The following reagent was obtained through the NIH HIV Reagent Program, Division of AIDS, NIAID, NIH: Human Immunodeficiency Virus 1 (HIV-1) NL4-3 ΔEnv Vpr Luciferase Reporter Vector (pNL4-3.Luc.R-E-), ARP-3418, contributed by Dr. Nathaniel Landau and Aaron Diamond. We thank our colleagues at both private and public testing laboratories, who submit samples for SARS-CoV-2 sequencing despite numerous challenges. We would like to acknowledge the teams within the National Institute for Communicable Diseases Centre for Respiratory Diseases and Meningitis, Sequencing Core Facility and Centre for HIV and STIs. We thank Hyrax Biosciences for the use of their Exatype platform, Bridge-the-Gap and the Cape Town Immunology Laboratory. In addition we would like to thank the originating and submitting laboratories (listed in **Supplementary Table 2**) for providing information regarding travel history of International C.1.2 samples. The Network for Genomic Surveillance South Africa (NGS-SA) is supported by the Strategic Health Innovation Partnerships Unit of the South African Medical Research Council, with funds received from the South African Department of Science and Innovation. Sequencing activities for the different sequencing hubs were provided by a conditional grant from the South African National Department of Health as part of the emergency COVID-19 response, a cooperative agreement between the National Institute for Communicable Diseases of the National Health Laboratory Service and the United States Centers for Disease Control and Prevention (grant number 5 U01IP001048-05-00); the African Society of Laboratory Medicine (ASLM) and Africa Centers for Disease Control and Prevention through a sub-award from the Bill and Melinda Gates Foundation grant number INV-018978; the UK Foreign, Commonwealth and Development Office and Wellcome (Grant no 221003/Z/20/Z); the South African Medical Research Council (Reference number SHIPNCD 76756); the UK Department of Health and Social Care and managed by the Fleming Fund and performed under the auspices of the SEQAFRICA project; German Federal Ministry of Education and Research (BMBF; grant number 01KA1606; and G7 collaboration grant with the Robert Koch Institute for COVID19) for the African Network for Improved Diagnostics, Epidemiology and Management of common infectious Agents (ANDEMIA). This study was supported by the Bill and Melinda Gates award INV-018944 (AS), National Institutes of Health award R01 AI138546 (AS), South African Medical Research Council awards (AS, TdO, PLM) and National Institutes of Health U01 AI151698 for the United World Antivirus Research Network (UWARN) (WVV). Hyrax Biosciences’ Exatype platform was supported by the South African Medical Research Council with funds received from the Department of Science and Innovation. The content and findings reported/ illustrated are the sole deduction, view and responsibility of the researcher and do not reflect the official position and sentiments of the SAMRC or the Department of Science and Innovation. SIR is a L’Oreal/UNESCO Women in Science South African Young Talents awardee. DPM and CW were supported by the Wellcome Trust (222574/Z/21/Z). CW and JNB are funded by the EDCTP (RADIATES Consortium; RIA2020EF-3030). PLM is supported by the South African Research Chairs Initiative of the Department of Science and Innovation and the NRF (Grant No 98341) and the Strategic Health Innovations Program of the SA MRC. The funders had no role in study design, data collection and analysis, decision to publish, or preparation of the manuscript.

## Author Contributions

CS and JE: designed the study, identified the lineage, performed sequence analysis and wrote the manuscript; DGA, HT, DPM, EW, JES performed sequence analysis; CKW performed the modelling of mutations within the SARS-CoV-2 spike; AM, AI, BM, JG, NN, SP, TM, UR, YN and ZTK performed SARS-CoV-2 sequencing; BEL made plasmids encoding the SARS-CoV-2 C.1.2 spike; SIR designed ADCC experiments and provided vaccination samples and NM performed the ADCC experiments; PK and SC performed the neutralization assays; GK, GG, L-GB, SAM and WVV provided vaccination samples; VB provided vaccine breakthrough samples for SARS-CoV-2 sequencing; NGS-SA is a consortium responsible for SARS-CoV-2 sequencing and analysis for the genomic surveillance of SARS-CoV-2 within South Africa; FT and MV co-ordinate specimen collection for SARS-CoV-2 sequencing, NW and RL performed epidemiological analyses; KM is an NGS-SA PI and co-ordinates SARS-CoV-2 specimen submission for sequencing and responsible for source funding; CW, N-YH, NM, TM, WP, ZM, RL, TdO and AvG are NGS-SA Principal Investigator (PI)s and co-ordinate SARS-CoV-2 sequencing and responsible for source funding; AS designed neutralization experiments; PLM designed neutralization experiments and wrote the manuscript; JNB designed study, performed sequence analysis and wrote the manuscript. All authors commented on the manuscript.

## METHODS

Methods and any associated references are available in the online version of the paper.

### Data Availability

All SARS-CoV-2 assemblies used in this analysis are deposited in GISAID (https://www.gisaid.org/)^21,22^ and the GISAID accessions are provided in **Supplementary Tables 1** and **2**. The Nextstrain build of C.1.2 and global sequences will be made available at https://nextstrain.org/groups/ngs-sa. The alignment used for the recombination analysis is provided as an extended data file on the online version of the paper.

## SUPPLEMENTAL INFORMATION

## ONLINE METHODS

### Samples and Ethics Approvals

#### SARS-CoV-2 Genomic Surveillance

As part of monitoring the viral evolution by the Network for Genomics Surveillance of South Africa (NGS-SA)^5^, seven sequencing hubs receive randomly selected samples for sequencing every week according to approved protocols at each site. These samples include remnant nucleic acid extracts or remnant nasopharyngeal and oropharyngeal swab samples from routine diagnostic SARS-CoV-2 PCR testing, from public and private laboratories in South Africa. Permission was obtained for associated metadata for the samples including date and location (district and province) of sampling, and sex and age of the patients to offer additional insights about the epidemiology of the infection caused by the virus. The project was approved by the University of the Witwatersrand Human Research Ethics Committee (HREC) (ref. M180832, M210159, M210752), University of KwaZulu–Natal Biomedical Research Ethics Committee (ref. BREC/00001510/2020), Stellenbosch University HREC (ref. N20/04/008_COVID19) and the University of Cape Town HREC (ref. 383/2020) and the University of Pretoria, Faculty of Health human ethics committee, (ref H101-2017). Individual participant consent was not required for the genomic surveillance. This requirement was waived by the Research Ethics Committees.

#### Hospitalized Steve Biko Cohort

This study has been given ethics approval by the University of Pretoria, Human Research Ethics Committee (Medical) (247/2020). Serum samples were obtained (longitudinally) from hospitalized patients with PCR-confirmed SARS-CoV-2 infection, known HIV status and aged ≥18 years. These samples have previously been used to assess antibody responses to the D614G and Beta variants^12^, a subset of seven of these were used to measure wave 1 and wave 3 immune responses to Delta and C.1.2 using the pseudovirus neutralization assay and ADCC activity against D614G and C.1.2. These samples had a median of 3 days post-PCR test.

#### Groote Schuur Hospital Cohort

Plasma samples were obtained from hospitalized COVID-19 patients with moderate disease admitted to Groote Schuur Hospital cohort, Cape Town from 30 December 2020 – 15 January 2021 during the second wave in South Africa. All patients were aged ≥18 years and were HIV negative. This study received ethics approval from the Human Research Ethics Committee of the Faculty of Health Sciences, University of Cape Town (R021/2020). Neutralization activity has previously been assessed for this cohort against the D614G and Beta variants^23^, a subset of seven of these were used to measure wave 2 antibody responses against Delta and C.1.2 using the pseudovirus neutralization assay and ADCC activity against D614G and C.1.2.

#### ChAdOx1 nCOV-19 (AZD1222) Vaccinees’ Samples

Samples from donors vaccinated with the ChAdOx1 nCOV-19 (AZD1222) vaccine were previously assessed for neutralization activity against the D614G and Beta variants^24^. This study received ethics approval from the Pan African Clinical Trials Registry (PACTR202006922165132) as well as the South Africa Health Products Regulatory Authority (SAHPRA: 20200407). A subset of eleven of these samples were used to test neutralization activity against Delta and C.1.2 using the pseudovirus neutralization assay and ADCC activity against D614G and C.1.2.

#### Janssen/Johnson and Johnson (Ad26.COV2.S) Vaccinees’ Samples

nine samples from healthy donors vaccinated with the Janssen/Johnson and Johnson (Ad26.COV2.S) vaccine during the Sisonke Trial were obtained 2 months after vaccination and used for the pseudovirus neutralization against D614G, Beta, Delta and C.1.2 and ADCC activity against D614G and C.1.2. Ethics approval for the use of these samples were obtained from the Human Research Ethics Committee of the Faculty of Health Sciences, University of the Witwatersrand (M210465).

#### Pfizer/BioNTech (BNT162b2) Vaccinees’ Samples

Seven samples from donors vaccinated with BNT162b2 were used for the pseudovirus neutralization assay against D614G, Beta, Delta and C.1.2 and ADCC activity against D614G and C.1.2. These samples were obtained from healthy donors two months after their second dose. Ethics approval for the use of these samples were obtained from the Human Research Ethics Committee of the Faculty of Health Sciences, University of the Witwatersrand (M210465). For the live virus neutralization assay, six samples were collected and provided by the United World Antivirus Research Network (UWARN), participants were between the ages of 20 - 59 and samples were collected from 2.9 to 5.1 months post second vaccination dose. Ethics approval for the use of these samples was obtained by the Biomedical Research Ethics Committee at the University of KwaZulu–Natal (BREC/00001275/2020).

### SARS-CoV-2 Whole-Genome Sequencing and Genome Assembly

#### RNA Extraction

RNA was extracted either manually or automatically in batches, using the QIAamp viral RNA mini kit (QIAGEN, California, USA) as per manufacturer’s instructions, using the Nucleo Mag Pathogen kit (Macherey-Nagel, Duren, Germany) on a Hamilton Microlab STAR instrument (Hamilton Company, Reno, NV) or the Chemagic 360 using the CMG-1049 kit (PerkinElmer, Massachusetts, USA). A modification was done on the manual extractions by adding 280 µl per sample, in order to increase yields. 300 µl of each sample was used for automated magnetic bead-based extraction using the Chemagic 360. RNA was eluted in 60 µl of the elution buffer. Isolated RNA was stored at -80°C prior to use.

### PCR and Library Preparation

Sequencing was performed using the Illumina COVIDSeq protocol (Illumina Inc, USA) or nCoV-2019 ARTIC network sequencing protocol v3 (https://artic.network/ncov-2019). These are amplicon-based next-generation sequencing approaches. Briefly, for nCoV-2019 ARTIC network sequencing protocol, the first strand synthesis was carried out on extracted RNA samples using random hexamers primers from the SuperScript IV reverse transcriptase synthesis kit (Life Technologies) or LunaScript RT SuperMix Kit (New England Biolabs (NEB), Ipswich, MA). The synthesized cDNA was amplified using multiplex polymerase chain reactions (PCRs) using ARTIC nCoV-2019 v3 primers. For COVIDSeq protocol, the first strand synthesis was carried using random hexamers primers from Illumina and the synthesized cDNA underwent two separate multiplex PCR reactions.

*For Illumina sequencing* using the nCoV-2019 ARTIC network sequencing protocol, the pooled PCR products underwent bead-based tagmentation using the Nextera Flex DNA library preparation kit. The adapter-tagged amplicons were cleaned-up using AmpureXP purification beads (Beckman Coulter, High Wycombe, U and amplified using one round of PCR. The PCRs were indexed using the Nextera CD indexes (Illumina, San Diego, CA, USA) according to the manufacturer’s instructions. For COVIDSeq sequencing protocol, pooled PCR amplified products were processed for tagmentation and adapter ligation using IDT for Illumina Nextera UD Indexes. Further enrichment and cleanup was performed as per protocols provided by the manufacturer (Illumina Inc). Pooled samples from both COVIDSeq protocol and nCoV-2019 ARTIC network protocol were quantified using Qubit 3.0 or 4.0 fluorometer (Invitrogen Inc.) using the Qubit dsDNA High Sensitivity assay according to manufacturer’s instructions. The fragment sizes were analyzed using TapeStation 4200 (Invitrogen). The pooled libraries were further normalized to 4nM concentration and 25 μl of each normalized pool containing unique index adapter sets were combined in a new tube. The final library pool was denatured and neutralized with 0.2N sodium hydroxide and 200 mM Tris-HCL (pH7), respectively. 1.5 pM sample library was spiked with 2% PhiX. Libraries were loaded onto a 300-cycle NextSeq 500/550 HighOutput Kit v2 and run on the Illumina NextSeq 550 instrument (Illumina, San Diego, CA, USA).

#### For Oxford Nanopore sequencing

PCR products were quantified, without prior cleaning, using the Qubit dsDNA High Sensitivity assay on the Qubit 2.0 fluorometer (Thermo Fisher Scientific, Waltham, MA). Following DNA repair (NEB) and end-prep reactions (NEB), up to 96 samples were barcoded by ligation using the EXP-NBD196 kit (Oxford Nanopore Technologies (ONT), Oxford, U.K.). Barcoded samples were pooled, bead-purified and ligated to sequencing adapters using the Adapter Mix II Expansion kit (ONT). After the bead-purification, the DNA concentration was quantified on the Qubit 2.0 instrument (Thermo Fisher). Up to 100 ng of the library was diluted in 75 μl of sequencing mix loaded on an R9 flow-cell (ONT). A sequencing experiment was performed using MinKNOW software on the GridION X5 (ONT), with the high-accuracy base-calling setting. The NC045512 reference was used for alignment during base-calling and the barcodes were split into different folders

##### Assembly, Processing and Quality Control of Genomic Sequences

Raw reads from Illumina sequencing were assembled using the Exatype NGS SARS-CoV-2 pipeline v1.6.1, (https://sars-cov-2.exatype.com/) or Genome Detective 1.132/1.133 (https://www.genomedetective.com/) and the Coronavirus Typing Tool^25,26^. Samples sequenced from Oxford Nanopore GridION were assembled according to the Arctic-nCoV2019 novel coronavirus bioinformatics protocol or using the Fastq QC + ARTIC + NextClade pipeline on Epi2Me (Oxford Nanopore Technologies). For these samples raw reads were base called and demultiplexed using Guppy. To guarantee accuracy of the base calls, we only used dual indexed reads (i.e. required barcodes at both ends). A reference-based assembly and mapping was generated for each sample using Minimap2 and consensus calculated using Nanopolish. The reference genome used throughout the assembly process was NC_045512.2 (Accession number: MN908947.3). The initial assembly obtained was cleaned by aligning mapped reads to the references and filtering out low-quality mutations using the Geneious software v2021.0.3 (Biomatters). Quality control reports were obtained from Nextclade^27^. The resulting consensus sequence was further manually polished by considering and correcting indels in homopolymer regions that break the open reading frame (probably sequencing errors) using Aliview v1.27, (http://ormbunkar.se/aliview/)^28^. Mutations resulting in mid-gene stop codons and frameshifts were reverted to wild type. Regions with clustered mutations and deletions resulting in frameshifts were annotated as gaps and insertions were removed. Sequences with less than 80% coverage relative to the Wuhan-Hu-1 reference were discarded. All assemblies were deposited in GISAID (https://www.gisaid.org/)^21^ and the GISAID accession was included as part of **Supplementary Table 1**. Clade and lineage assignment was determined using Nextclade and Pangolin^29^.

### Classification of Lineage, Clade and Associated Mutations

The ‘Phylogenetic Assignment of Named Global Outbreak Lineages’ (PANGOLIN) software suite (https://github.com/hCoV-2019/pangolin) was used for the dynamic SARS-CoV-2 lineage classification^29^. The SARS-CoV-2 genomes in our dataset were also classified using the clade classification proposed by NextStrain (https://nextstrain.org/) built for real-time tracking of the pathogen evolution^30^. The PANGO lineage identified predominantly in South Africa in this study is now assigned to the lineage C.1.2 (Pangolin version v3.1.7, lineages version 2021-07-28); the corresponding Nextclade classification is 20D (Nextclade version v1.5.3, clades version 2021-07-28). The C.1.2 lineage and its associated mutations were further confirmed using the Stanford Coronavirus Antiviral & Resistance Database (CoVDB) (https://covdb.stanford.edu/) and Outbreak.info (https://outbreak.info/).

### Dataset Compilation

At the time of writing, there were over 3.7 million SARS-CoV-2 genomes available on GISAID (https://www.gisaid.org). Due to the size of this dataset, sub-sampling was performed to obtain a representative but manageable sample of genomes. A preliminary dataset was downloaded from GISAID. For C.1.2, genomes were manually curated to include all genomes with more than 90% coverage and complete date information. Due to the dynamic nature of GISAID submissions we did not include sequences that were released later than September 10th, with the exception of the Eswatini sequences (**Supplementary table 2**) which we had sequenced and deposited on GISAID (with permission of the originating laboratory) and so could access before their release date. Following this, C.1 (the original lineage to which C.1.2 was assigned), C.1.1 (a Mozambican lineage that evolved from C.1^7^) and South African genomes were downloaded with the options ‘complete’, ‘low coverage excluded’, and ‘collection date complete’ selected to ensure that only genomes with complete date information and more than 95% coverage were included. The global and African Auspice datasets were also downloaded (accessed 10 September 2021). This dataset was further down-sampled using a custom build of the Nextstrain SARS-CoV-2 pipeline^30^ to produce a final dataset of 6,192 genomes. Of these, 156 are from lineage C.1.2. Due to the fact that C.1.2 was first detected and is most prevalent in South Africa, we chose to include a large proportion of South African sequences, resulting in 1,991 South African genomes. To include global context, there were an additional 1,047 sequences from the rest of Africa, 902 from Asia, 1,132 from Europe, 484 from South America, 430 from North America, and 206 from Oceania. This dataset included genomes from all Variants of Concern (VOC) and Variants of Interest (VOI) as defined by the WHO at the time of writing (www.who.int, accessed 17 September 2021).

### Temporal Analysis

Using a dataset of global C.1.2 sequences publicly available on GISAID, we constructed a maximum-likelihood tree in IQ-tree^31^. We inspected this maximum-likelihood tree in TempEst v.1.5.3 for the presence of a temporal (that is, molecular clock) signal^32^. The regression of root-to-tip distance against sampling date showed that the sequences evolved in a relatively strong clock-like manner, with a correlation coefficient of 0.43 and R^2^ of 0.18.

### Phylogenetic Analysis and Recombination Detection

Phylogenetic analysis was conducted with a custom Nextstrain SARS-CoV-2 build^30^. Briefly, the pipeline filters sequences, aligns these sequences with Nextalign (https://github.com/neherlab/nextalign), sub-samples the datasets (resulting in the dataset described above), constructs a phylogenetic tree with IQ-TREE^31^, refines and dates the tree with TreeTime^33^, reconstructs ancestral states, and assigns Nextstrain clades to the sequences. The tree was visualized with Auspice to confirm the presence of a C.1.2 cluster. This revealed that several non-C.1.2 samples clustered with C.1.2. These sequences were inspected for the presence of the major C.1.2 mutations (dark purple mutations in **Fig. 3b**). All sequences possessed a subset of the major mutations (a number of sequences had missing data spanning the regions of defining mutations); this, along with the clustering, was used as evidence to re-assign the sequences to C.1.2, resulting in a set of 156 C.1.2 genomes.

To model phylogenetic diffusion duplicate Markov Chain Monte Carlo (MCMC) analyses were executed in BEAST v1.10.4 for 100 million iterations with sampling every 10000 steps in the chain. For each sequence, latitude and longitude were attributed to a point randomly sampled within the local area or district of origin. For this analysis, we used the strict molecular clock model, the HKY+I, nucleotide substitution model and the exponential growth coalescent model^34^. Convergence of runs was assessed in Tracer v.1.7.1 based on high effective sample sizes and good mixing^35^. Maximum clade credibility trees for each run were summarized using TreeAnnotator after discarding the first 10% of the chain as burn-in. The R package “seraphim’’ was used to extract and map the spatiotemporal information embedded in the MCC trees^36^.

We tested for evidence of recombination with RDP5^37^ using seven recombination detection methods implemented therein and a liberal multiple testing uncorrected p-value cutoff of 0.05 to maximize power to detect recombination. Specifically, we screened an alignment of twelve representative C.1.2, eight Alpha sequences, eight Beta sequences, nine Delta sequences and six C.1 sequences for evidence that the mutations shared by the VOCs and C.1.2 had been acquired by recombination between viruses in the VOC and C.1 lineages. The specific sequences selected for analysis were representative of the diversity within their respective lineages in context of the South African epidemic.

### SARS-CoV-2 Model

We modelled the spike protein on the basis of the Protein Data Bank coordinate set 7A94. We used the Pymol program (The PyMOL Molecular Graphics System, version 2.2.0) for visualization.

### Lentiviral Pseudovirus Production and Neutralization Assay

Virus production and pseudovirus neutralization assays were done as previously described^12^. Briefly, 293T/ACE2.MF cells modified to overexpress human ACE2 (kindly provided by M. Farzan (Scripps Research)) were cultured in DMEM (Gibco BRL Life Technologies) containing 10% heat-inactivated serum (FBS) and 3 μg ml−1 puromycin at 37 °C, 5% CO_2_. Cell monolayers were disrupted at confluency by treatment with 0.25% trypsin in 1 mM EDTA (Gibco BRL Life Technologies). The SARS-CoV-2, Wuhan-1 spike, cloned into pCDNA3.1 was mutated using the QuikChange Lightning Site-Directed Mutagenesis kit (Agilent Technologies) and NEBuilder HiFi DNA Assembly Master Mix (NEB) to include D614G (wild-type) or lineage defining mutations for Beta (L18F, D80A, D215G, 241-243del, K417N, E484K, N501Y, D614G and A701V), Delta (T19R, 156-157del, R158G, L452R, T478K, D614G, P681R and D950N) and C.1.2 (P9L, C136F, Y144del, R190S, D215G, 242-243del, Y449H, E484K, N501Y, D614G, H655Y, N679K, T716I and T859N). Pseudoviruses were produced by co-transfection in 293T/17 cells with a lentiviral backbone (HIV-1 pNL4.luc encoding the firefly luciferase gene) and either of the SARS-CoV-2 spike plasmids with PEIMAX (Polysciences). Culture supernatants were clarified of cells by a 0.45-μM filter and stored at −70 °C. Plasma samples were heat-inactivated and clarified by centrifugation. Pseudovirus and serially diluted plasma/sera were incubated for 1 h at 37 °C, 5% CO_2_. Cells were added at 1 × 104 cells per well after 72 h of incubation at 37 °C, 5% CO_2_, luminescence was measured using PerkinElmer Life Sciences Model Victor X luminometer. Neutralization was measured as described by a reduction in luciferase gene expression after single-round infection of 293T/ACE2.MF cells with spike-pseudotyped viruses. Titers were calculated as the reciprocal plasma dilution (ID50) causing 50% reduction of relative light units.

### Viral Expansion and Live Virus Neutralization Assay

All work with live virus was performed in Biosafety Level 3 containment using protocols for SARS-CoV-2 approved by the Africa Health Research Institute Biosafety Committee. ACE2-expressing H1299-E3 cells were used for the initial isolation (P1 stock) followed by passaging in Vero E6 cells (P2 and P3 stocks, where P3 stock was used in experiments). ACE2-expressing H1299-E3 cells were seeded at 1.5 × 10^5^ cells per mL and incubated for 18–20 h. After one DPBS wash, the sub-confluent cell monolayer was inoculated with 500 μL universal transport medium diluted 1:1 with growth medium filtered through a 0.45-μm filter. Cells were incubated for 1 h. Wells were then filled with 3 mL complete growth medium. After 8 days of infection, cells were trypsinized, centrifuged at 300 rcf for 3 min and resuspended in 4 mL growth medium. Then 1 mL was added to Vero E6 cells that had been seeded at 2 × 10^5^ cells per mL 18–20 h earlier in a T25 flask (approximately 1:8 donor-to-target cell dilution ratio) for cell-to-cell infection. The coculture of ACE2-expressing H1299-E3 and Vero E6 cells was incubated for 1 h and the flask was then filled with 7 mL of complete growth medium and incubated for 6 days. The viral supernatant (P2 stock) was aliquoted and stored at −80 °C and further passaged in Vero E6 cells to obtain the P3 stock used in experiments as follows: a T25 flask (Corning) was seeded with Vero E6 cells at 2 × 10^5^ cells per mL and incubated for 18–20 h. After one DPBS wash, the sub-confluent cell monolayer was inoculated with 500 μL universal transport medium diluted 1:1 with growth medium and filtered through a 0.45-μm filter. Cells were incubated for 1 h. The flask was then filled with 7 mL of complete growth medium. After infection for 4 days, supernatants of the infected culture were collected, centrifuged at 300 rcf for 3 min to remove cell debris and filtered using a 0.45-μm filter. Viral supernatant was aliquoted and stored at −80 °C.

Vero E6 cells were plated in a 96-well plate (Corning) at 30,000 cells per well 1 day before infection. Approximately 5 mL sterile water was added between wells to prevent wells at the edge drying more rapidly, which we have observed to cause edge effects resulting in lower number of foci. Plasma was separated from EDTA-anticoagulated blood by centrifugation at 500 rcf for 10 min and stored at −80 °C. Aliquots of plasma samples were heat-inactivated at 56 °C for 30 min and clarified by centrifugation at 10,000 rcf for 5 min, after which the clear middle layer was used for experiments. Inactivated plasma was stored in single-use aliquots to prevent freeze–thaw cycles. For experiments, plasma was serially diluted two-fold from 1:100 to 1:1,600; this is the concentration that was used during the virus–plasma incubation step before addition to cells and during the adsorption step. As a positive control, the GenScript A02051 anti-spike monoclonal antibody was added. Virus stocks were used at approximately 50-100 focus-forming units per microwell and added to diluted plasma; antibody–virus mixtures were incubated for 1 h at 37 °C, 5% CO_2_. Cells were infected with 100 μL of the virus–antibody mixtures for 1 h, to allow adsorption of virus. Subsequently, 100 μL of a 1X RPMI 1640 (Sigma-Aldrich, R6504), 1.5% carboxymethylcellulose (Sigma-Aldrich, C4888) overlay was added to the wells without removing the inoculum. Cells were fixed at 18 h after infection using 4% paraformaldehyde (Sigma-Aldrich) for 20 min. For staining of foci, a rabbit anti-spike monoclonal antibody (BS-R2B12, GenScript A02058) was used at 0.5 μg/mL as the primary detection antibody. Antibody was resuspended in a permeabilization buffer containing 0.1% saponin (Sigma-Aldrich), 0.1% BSA (Sigma-Aldrich) and 0.05% Tween-20 (Sigma-Aldrich) in PBS. Plates were incubated with primary antibody overnight at 4 °C, then washed with wash buffer containing 0.05% Tween-20 in PBS. Secondary goat anti-rabbit horseradish peroxidase (Abcam ab205718) antibody was added at 1 μg/mL and incubated for 2 h at room temperature with shaking. The TrueBlue peroxidase substrate (SeraCare 5510-0030) was then added at 50 μL per well and incubated for 20 min at room temperature. Plates were then dried for 2 h and imaged using a Metamorph-controlled Nikon TiE motorized microscope with a 2X objective or ELISPOT instrument with built-in image analysis (C.T.L). For microscopy images, automated image analysis was performed using a custom script in MATLAB v.2019b (Mathworks), in which focus detection was automated and did not involve user curation. Plasma dilutions used were 1:10, 1:20, 1:40, 1:80, 1:160, 1:320, 1:640, 1:1280 for self-plasma and 1:25, 1:50, 1:100, 1:200, 1:400, 1:800, 1:1600 for all other plasma samples tested.

### Antibody-Dependent Cellular Cytotoxicity (ADCC) Assay

The ability of plasma antibodies to cross-link FcγRIIIa (CD16) and spike expressing cells was measured as a proxy for ADCC. HEK293T cells were transfected with 5μg of SARS-CoV-2 wild-type variant spike (D614G), Beta, Delta or C.1.2 spike plasmids using PEI-MAX 40,000 (Polysciences) and incubated for 2 days at 37°C. Expression of spike was confirmed by binding of CR3022 and P2B-2F6 and their detection by anti-IgG APC staining measured by flow cytometry. Subsequently, 1×105 spike transfected cells per well were incubated with heat inactivated plasma (1:100 final dilution) or control mAbs (final concentration of 100μg/ml) in RPMI 1640 media supplemented with 10% FBS 1% Pen/Strep (Gibco, Gaithersburg, MD) for 1 hour at 37°C. Jurkat-Lucia(tm) NFAT-CD16 cells (Invivogen) (2×105 cells/well) were added and incubated for 24 hours at 37°C, 5% CO2. Twenty μl of supernatant was then transferred to a white 96-well plate with 50μl of reconstituted QUANTI-Luc secreted luciferase and read immediately on a Victor 3 luminometer with 1s integration time. Relative light units (RLU) of a no antibody control were subtracted as background. Palivizumab was used as a negative control, while CR3022 was used as a positive control, and P2B-2F6 to differentiate the Beta from the D614G variant. To induce the transgene 1x cell stimulation cocktail (Thermofisher Scientific, Oslo, Norway) and 2 μg/ml ionomycin in R10 was added as a positive control.

### Statistical Analyses

A Wilcoxon matched pairs signed-rank test was used to measure the differences in neutralization ID_50_ titre between matched samples against the different viruses for both the live virus and pseudovirus neutralization assays, as well as the difference in ADCC RLU. P-values are reflected in the graphs, represented as an asterix where: * represents p<0.05, ** represents p<0.01, *** represents p<0.001 and **** represents p<0.0001.

### Data Availability

All of the global SARS-CoV-2 genomes of the C.1.2 lineage generated and presented in this article are publicly accessible through the GISAID platform (https://www.gisaid.org/), along with all other SARS-CoV-2 genomes generated by the NGS-SA. The GISAID accession identifiers of the C.1.2 sequences analyzed in this study are provided as part of **Supplementary Tables 1** and **2**, which also contain the metadata for the sequences. The nextstrain build of C.1.2 and global sequences will be made available at https://nextstrain.org/groups/ngs-sa.

**Supplementary Figure 1:**
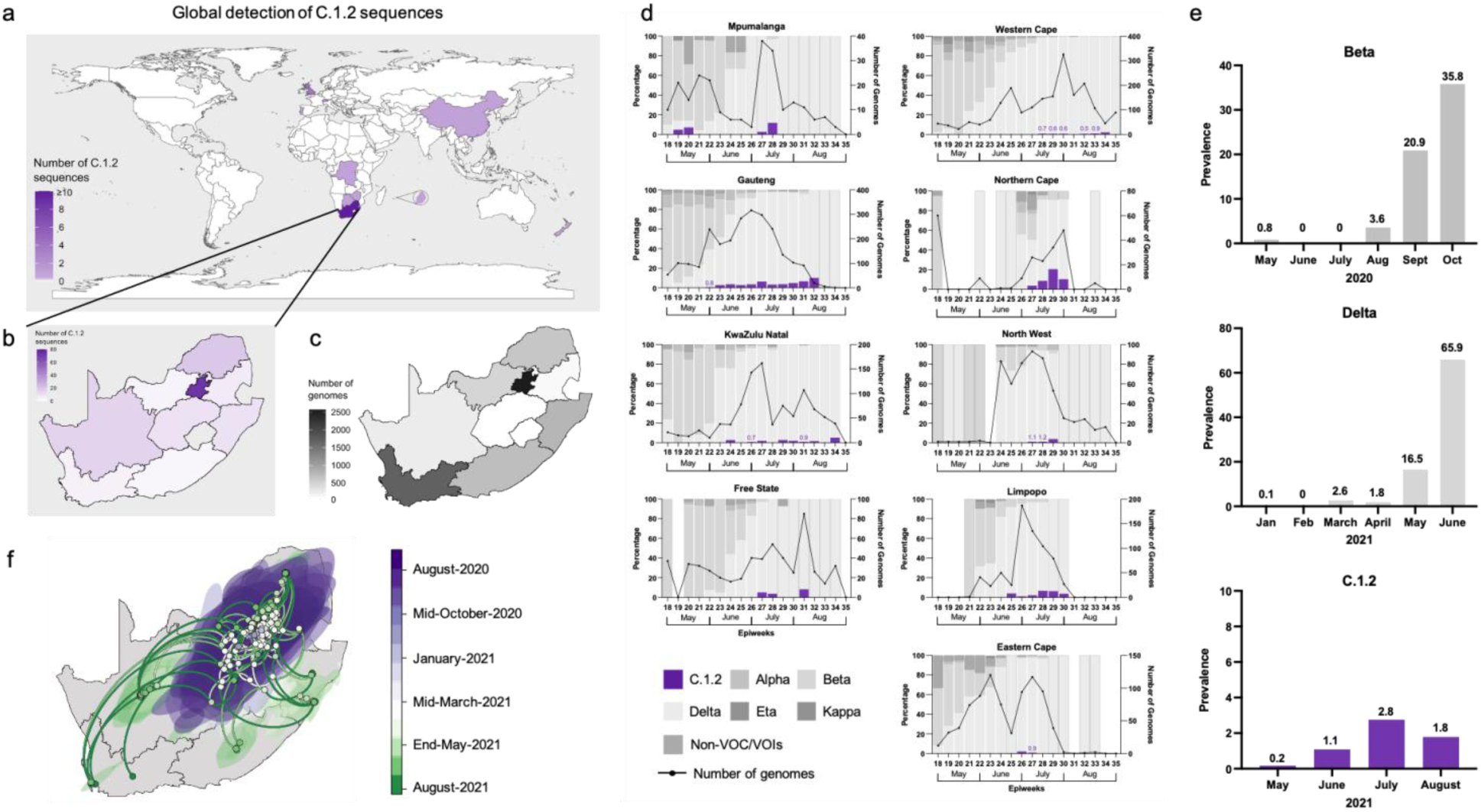
Distribution and prevalence of C.1.2 globally. **a**, Global map highlighting South Africa, Botswana, the Democratic Republic of the Congo, China, England, Eswatini, Mauritius (shown in the magnified bubble), New Zealand, Portugal, Switzerland and Zimbabwe, across which 166 C.1.2 sequences have been detected. Countries coloured according to the number of sequences detected. **b**, Map of South Africa highlighting the provinces in which C.1.2 has been detected, colored according to the number of sequences detected. **c**, Map of South Africa showing the number of SARS-CoV-2 genomes (n=8,337 as of September 10, 2021) that have been sequenced by province in the months of May to August 2021. **d**, Percentage of genomes that are assigned to various SARS-CoV-2 lineages by epidemiological week (epiweek) in South Africa for each of the provinces, with C.1.2 shown in purple (based on data submitted to GISAID on September, 21, 2021). The number of genomes sequenced for each epiweek is shown by the black line. **e**, Early prevalence rates of Beta, Delta and C.1.2 in South Africa based on the number of SARS-CoV-2 sequences generated for each month and submitted to GISAID on September 21, 2021. **f**, Spatiotemporal reconstruction of the spread of the C.1.2 cluster in South Africa during the third epidemic wave. Circles represent nodes of the maximum clade credibility phylogeny and are colored according to their inferred time of occurrence. Shaded areas represent the 80% highest posterior density interval and depict the uncertainty of the phylogeographic estimates for each node. Solid curved lines denote the links between nodes and the directionality of movement in an anticlockwise direction along the curve.

**Supplementary Figure 2:**
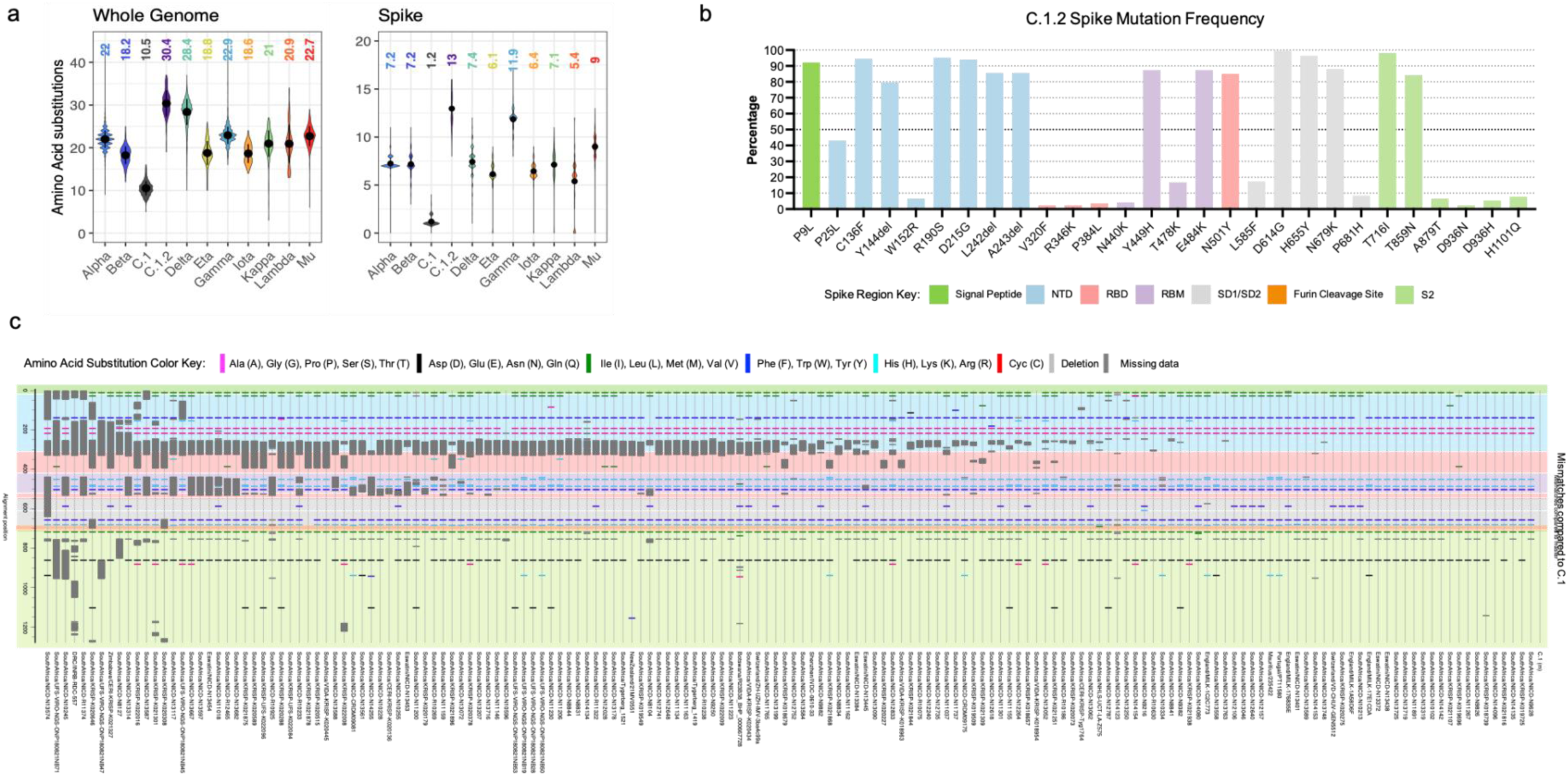
C.1.2 mutation profile. **a**, Whole genome (left panel) and spike region only (right panel) amino acid substitutions across different VOIs and VOCs compared to C.1 (the precursor of C.1.2) and C.1.2, coloured according to nextstrain clade, with C.1.2 in purple and C.1 in blue. The mean amino acid changes for each variant is shown in the plots. **b**, Frequency of spike mutations across C.1.2 sequences, though frequencies of these mutations may be underrepresented due to high levels of missing data shown in panel **c**. Regions within the spike are colored according to region as shown in the key. **c**, Highlighter plot of C.1.2 spike sequences with (n=166) identified across the globe, labelled according to the location identified and sequence name. Mismatches compared to the C.1 strain (top/master sequence) are colored by Se-Al in the hiv.lanl.gov highlighter tool as shown in the key. Large dark grey regions represent missing sequence data. Spike regions are also coloured according to the key in panel **b**.

**Supplementary Figure 3:**
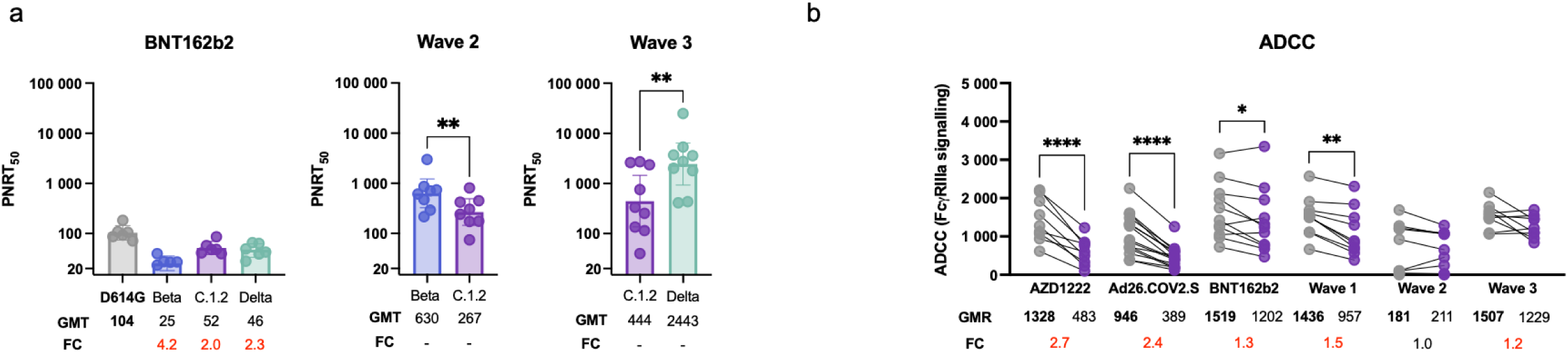
Live virus neutralization and ADCC profiles of C.1.2. **a**, Neutralization activity measure by the live virus neutralization assay from donors either vaccinated with BNT162b2 or previously infected during the second and third waves in South Africa against the wild-type (with D614G), Beta, Delta and C.1.2 variants. Bar graphs represent the geometric mean titer (GMT) for each group with the error bars representing the 95% confidence intervals, dots represent individual sample titers. Statistical significance based on the Wilcoxon test are shown above graphs. The * denotes p-values: * p<0.05 and ** p<0.01. GMT and fold-change (FC) differences relative to D614G (wild-type, shown in bold) are given below the graph, with red representing decrease in titre and green representing increases in titre. **b**, ADCC activity represented as relative light units (RLU) from plasma samples from donors either vaccinated with AZD1222, Ad26.COV.2.S or BNT162b2 or previously infected during waves one, two or three in South Africa against the wild-type (with D614G, shown as grey circles) and C.1.2 (shown as purple circles). P-values are denoted with “*” symbols: * p<0.05 and **** p<0.0001. Geometric mean of the RLU (GMR) and fold-changes (FC) relative to the wild-type (D614G, shown in bold) are given below the graph with red representing decreases in RLU and black representing no difference.

**Supplementary Table 1:**
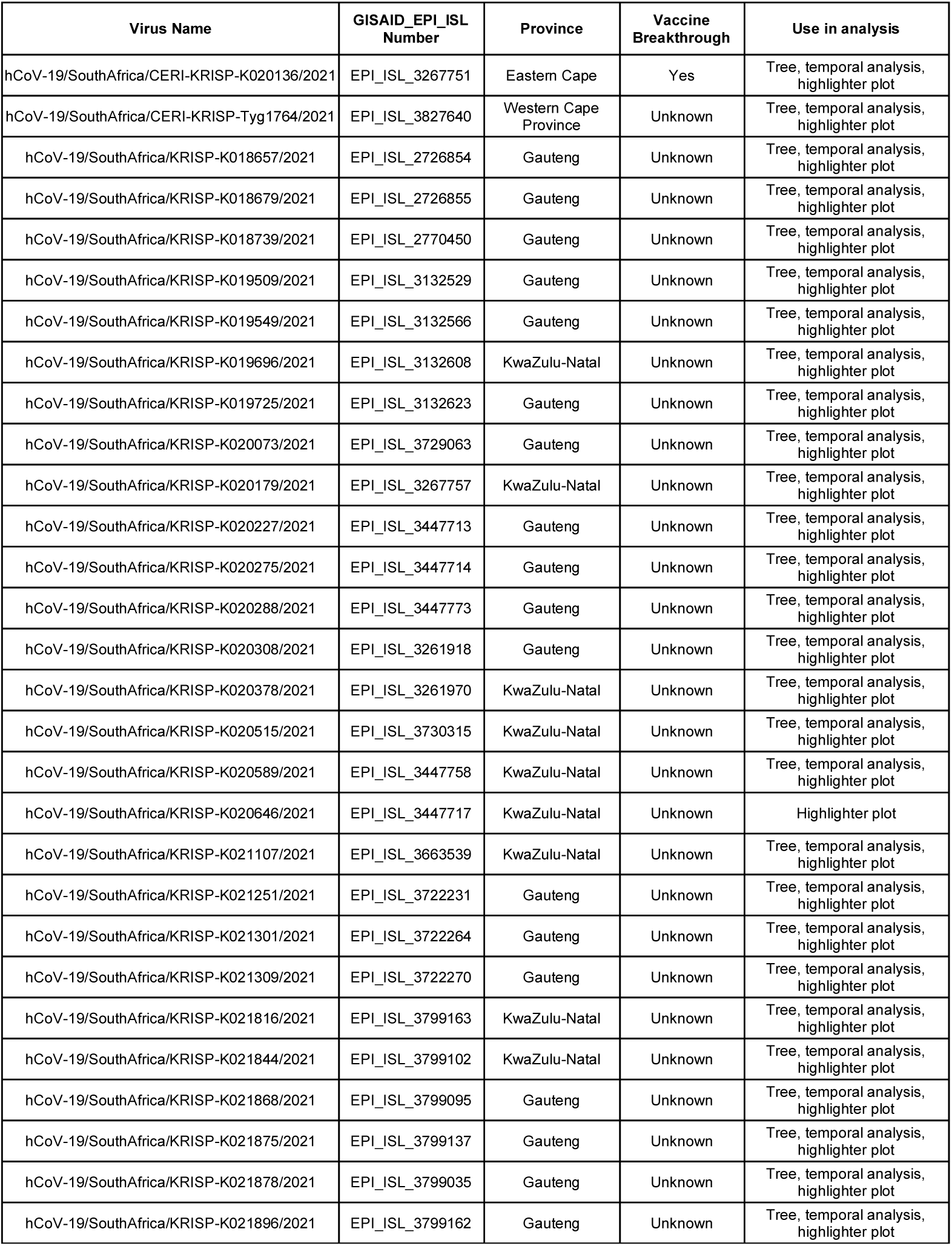
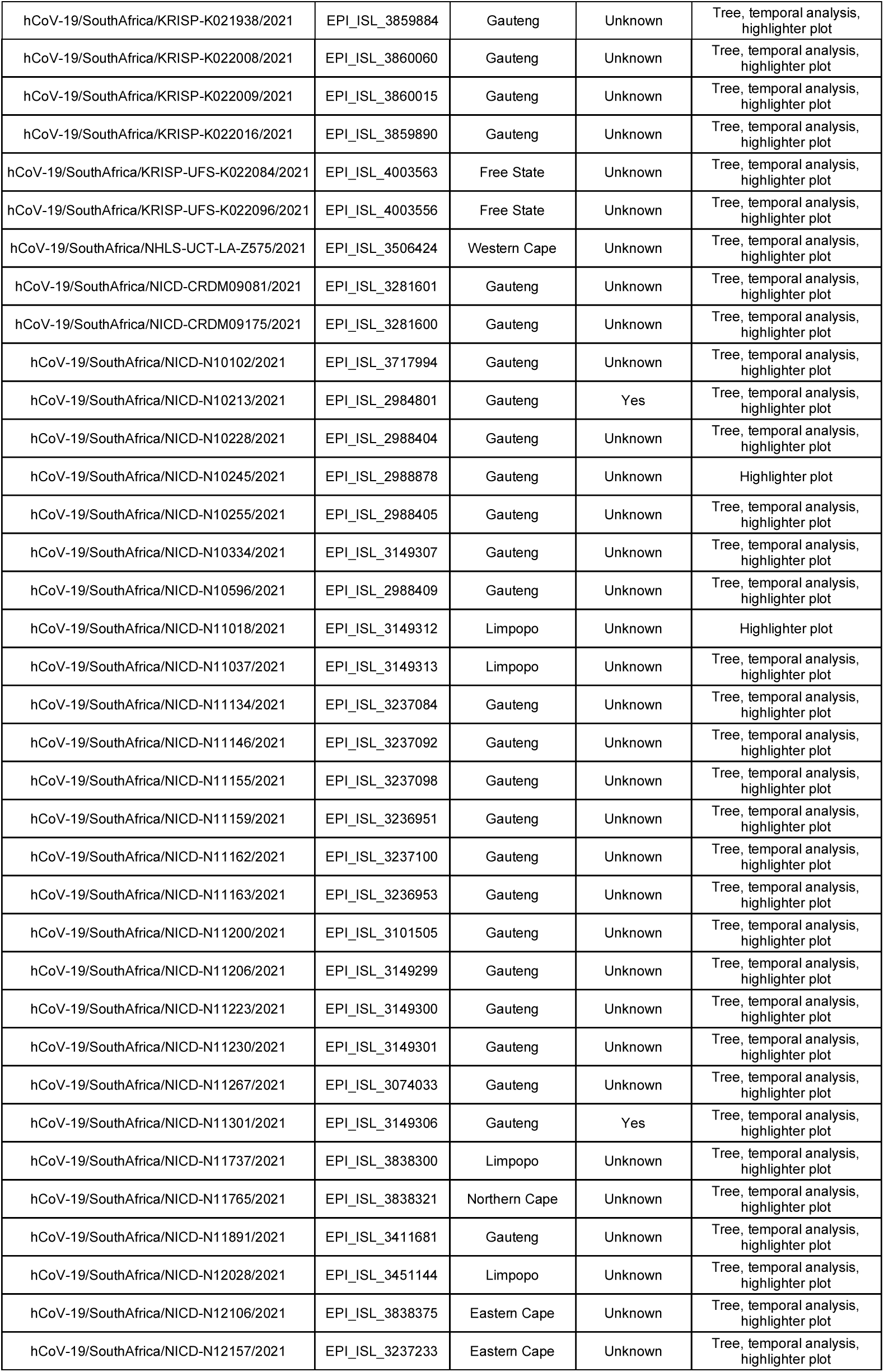
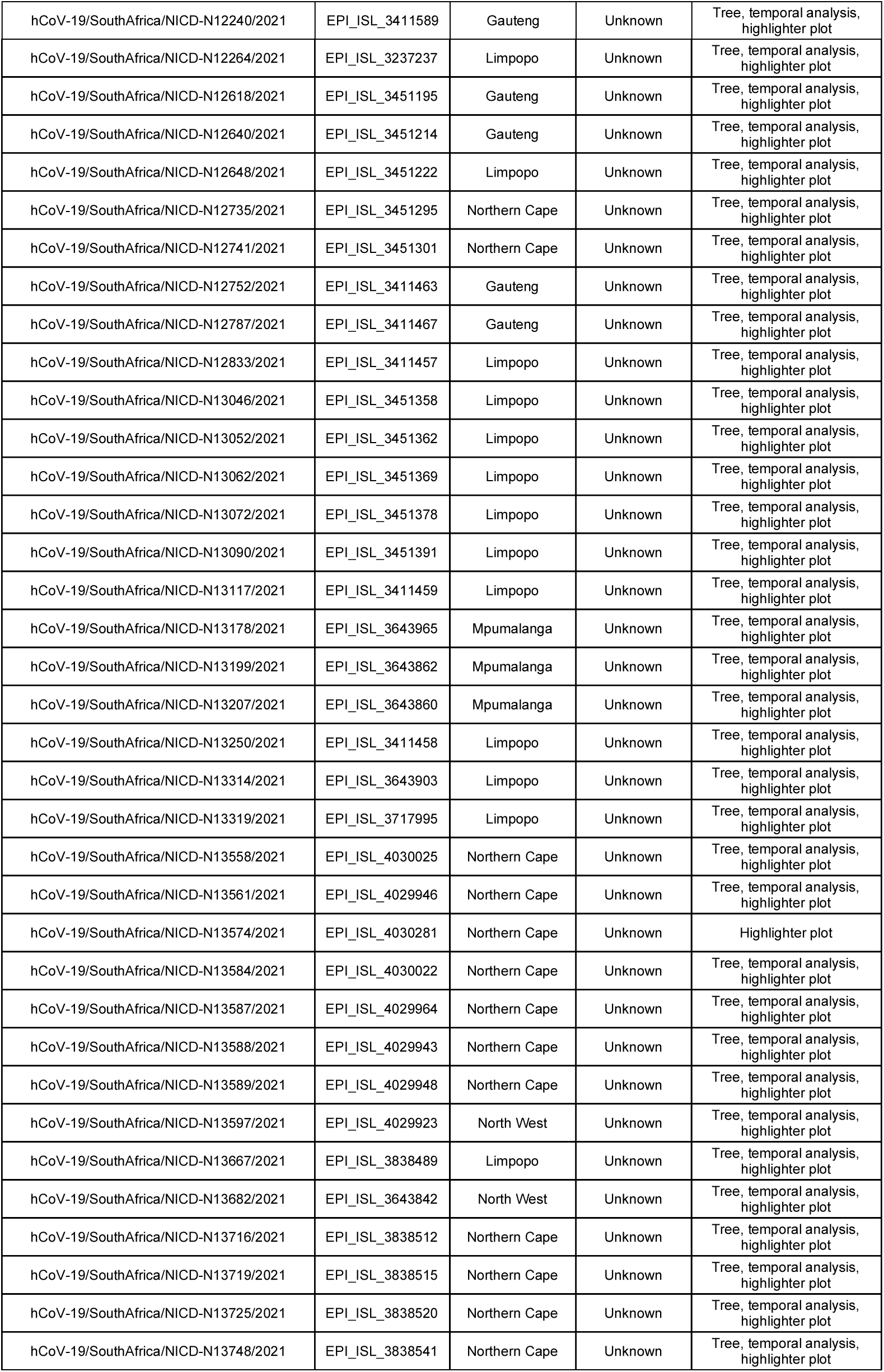
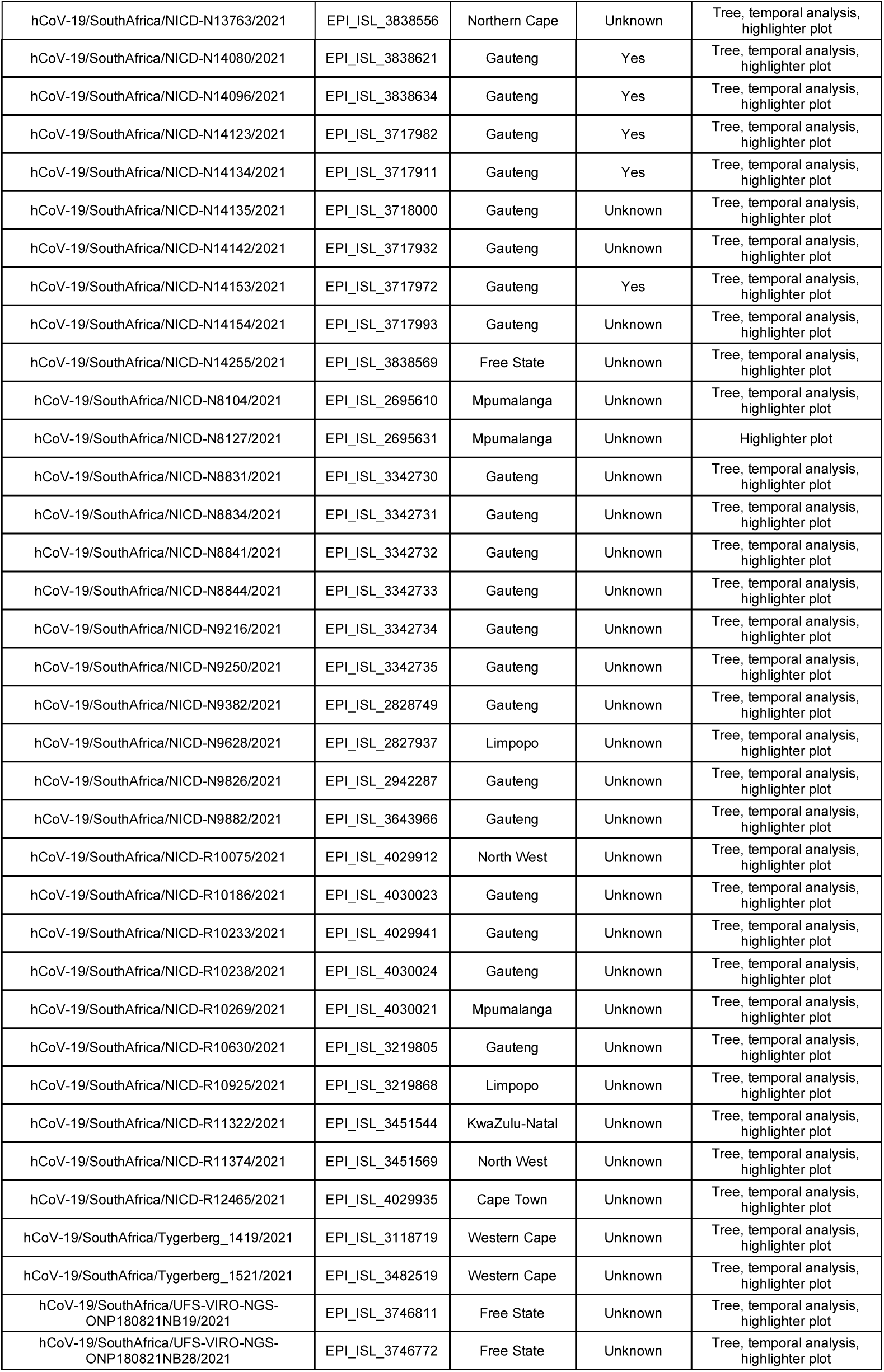
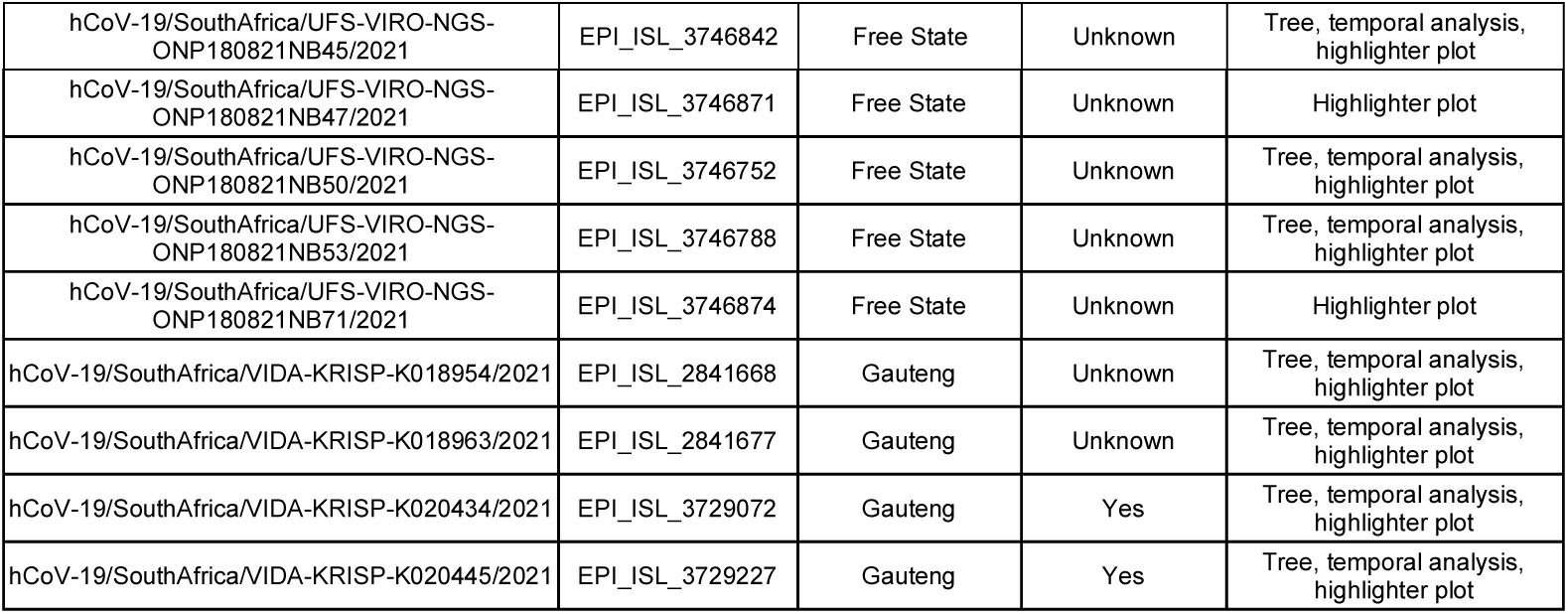
Reference set of C.1.2 genomes on GISAID from South Africa. Provided are the GISAID strain name and GISAID_EPI_ISL accession numbers for all C.1.2 sequences detected in South Africa as of data deposited on September 10, 2021. Provided are the provincial breakdown, vaccination breakthrough status and use in various analyses.

| Virus Name | GISAID_EPI_ISL Number | Province | Vaccine Breakthrough | Use in analysis |
| --- | --- | --- | --- | --- |
| hCoV-19/SouthAfrica/CERI-KRISP-K020136/2021 | EPI_ISL_3267751 | Eastern Cape | Yes | Tree, temporal analysis, highlighter plot |
| hCoV-19/SouthAfrica/CERI-KRISP-Tyg1764/2021 | EPI_ISL_3827640 | Western Cape Province | Unknown | Tree, temporal analysis, highlighter plot |
| hCoV-19/SouthAfrica/KRISP-K018657/2021 | EPI_ISL_2726854 | Gauteng | Unknown | Tree, temporal analysis, highlighter plot |
| hCoV-19/SouthAfrica/KRISP-K018679/2021 | EPI_ISL_2726855 | Gauteng | Unknown | Tree, temporal analysis, highlighter plot |
| hCoV-19/SouthAfrica/KRISP-K018739/2021 | EPI_ISL_2770450 | Gauteng | Unknown | Tree, temporal analysis, highlighter plot |
| hCoV-19/SouthAfrica/KRISP-K019509/2021 | EPI_ISL_3132529 | Gauteng | Unknown | Tree, temporal analysis, highlighter plot |
| hCoV-19/SouthAfrica/KRISP-K019549/2021 | EPI_ISL_3132566 | Gauteng | Unknown | Tree, temporal analysis, highlighter plot |
| hCoV-19/SouthAfrica/KRISP-K019696/2021 | EPI_ISL_3132608 | KwaZulu-Natal | Unknown | Tree, temporal analysis, highlighter plot |
| hCoV-19/SouthAfrica/KRISP-K019725/2021 | EPI_ISL_3132623 | Gauteng | Unknown | Tree, temporal analysis, highlighter plot |
| hCoV-19/SouthAfrica/KRISP-K020073/2021 | EPI_ISL_3729063 | Gauteng | Unknown | Tree, temporal analysis, highlighter plot |
| hCoV-19/SouthAfrica/KRISP-K020179/2021 | EPI_ISL_3267757 | KwaZulu-Natal | Unknown | Tree, temporal analysis, highlighter plot |
| hCoV-19/SouthAfrica/KRISP-K020227/2021 | EPI_ISL_3447713 | Gauteng | Unknown | Tree, temporal analysis, highlighter plot |
| hCoV-19/SouthAfrica/KRISP-K020275/2021 | EPI_ISL_3447714 | Gauteng | Unknown | Tree, temporal analysis, highlighter plot |
| hCoV-19/SouthAfrica/KRISP-K020288/2021 | EPI_ISL_3447773 | Gauteng | Unknown | Tree, temporal analysis, highlighter plot |
| hCoV-19/SouthAfrica/KRISP-K020308/2021 | EPI_ISL_3261918 | Gauteng | Unknown | Tree, temporal analysis, highlighter plot |
| hCoV-19/SouthAfrica/KRISP-K020378/2021 | EPI_ISL_3261970 | KwaZulu-Natal | Unknown | Tree, temporal analysis, highlighter plot |
| hCoV-19/SouthAfrica/KRISP-K020515/2021 | EPI_ISL_3730315 | KwaZulu-Natal | Unknown | Tree, temporal analysis, highlighter plot |
| hCoV-19/SouthAfrica/KRISP-K020589/2021 | EPI_ISL_3447758 | KwaZulu-Natal | Unknown | Tree, temporal analysis, highlighter plot |
| hCoV-19/SouthAfrica/KRISP-K020646/2021 | EPI_ISL_3447717 | KwaZulu-Natal | Unknown | Highlighter plot |
| hCoV-19/SouthAfrica/KRISP-K021107/2021 | EPI_ISL_3663539 | KwaZulu-Natal | Unknown | Tree, temporal analysis, highlighter plot |
| hCoV-19/SouthAfrica/KRISP-K021251/2021 | EPI_ISL_3722231 | Gauteng | Unknown | Tree, temporal analysis, highlighter plot |
| hCoV-19/SouthAfrica/KRISP-K021301/2021 | EPI_ISL_3722264 | Gauteng | Unknown | Tree, temporal analysis, highlighter plot |
| hCoV-19/SouthAfrica/KRISP-K021309/2021 | EPI_ISL_3722270 | Gauteng | Unknown | Tree, temporal analysis, highlighter plot |
| hCoV-19/SouthAfrica/KRISP-K021816/2021 | EPI_ISL_3799163 | KwaZulu-Natal | Unknown | Tree, temporal analysis, highlighter plot |
| hCoV-19/SouthAfrica/KRISP-K021844/2021 | EPI_ISL_3799102 | KwaZulu-Natal | Unknown | Tree, temporal analysis, highlighter plot |
| hCoV-19/SouthAfrica/KRISP-K021868/2021 | EPI_ISL_3799095 | Gauteng | Unknown | Tree, temporal analysis, highlighter plot |
| hCoV-19/SouthAfrica/KRISP-K021875/2021 | EPI_ISL_3799137 | Gauteng | Unknown | Tree, temporal analysis, highlighter plot |
| hCoV-19/SouthAfrica/KRISP-K021878/2021 | EPI_ISL_3799035 | Gauteng | Unknown | Tree, temporal analysis, highlighter plot |
| hCoV-19/SouthAfrica/KRISP-K021896/2021 | EPI_ISL_3799162 | Gauteng | Unknown | Tree, temporal analysis, highlighter plot |
| hCoV-19/SouthAfrica/KRISP-K021938/2021 | EPI_ISL_3859884 | Gauteng | Unknown | Tree, temporal analysis, highlighter plot |
| hCoV-19/SouthAfrica/KRISP-K022008/2021 | EPI_ISL_3860060 | Gauteng | Unknown | Tree, temporal analysis, highlighter plot |
| hCoV-19/SouthAfrica/KRISP-K022009/2021 | EPI_ISL_3860015 | Gauteng | Unknown | Tree, temporal analysis, highlighter plot |
| hCoV-19/SouthAfrica/KRISP-K022016/2021 | EPI_ISL_3859890 | Gauteng | Unknown | Tree, temporal analysis, highlighter plot |
| hCoV-19/SouthAfrica/KRISP-UFS-K022084/2021 | EPI_ISL_4003563 | Free State | Unknown | Tree, temporal analysis, highlighter plot |
| hCoV-19/SouthAfrica/KRISP-UFS-K022096/2021 | EPI_ISL_4003556 | Free State | Unknown | Tree, temporal analysis, highlighter plot |
| hCoV-19/SouthAfrica/NHLS-UCT-LA-Z575/2021 | EPI_ISL_3506424 | Western Cape | Unknown | Tree, temporal analysis, highlighter plot |
| hCoV-19/SouthAfrica/NICD-CRDM09081/2021 | EPI_ISL_3281601 | Gauteng | Unknown | Tree, temporal analysis, highlighter plot |
| hCoV-19/SouthAfrica/NICD-CRDM09175/2021 | EPI_ISL_3281600 | Gauteng | Unknown | Tree, temporal analysis, highlighter plot |
| hCoV-19/SouthAfrica/NICD-N10102/2021 | EPI_ISL_3717994 | Gauteng | Unknown | Tree, temporal analysis, highlighter plot |
| hCoV-19/SouthAfrica/NICD-N10213/2021 | EPI_ISL_2984801 | Gauteng | Yes | Tree, temporal analysis, highlighter plot |
| hCoV-19/SouthAfrica/NICD-N10228/2021 | EPI_ISL_2988404 | Gauteng | Unknown | Tree, temporal analysis, highlighter plot |
| hCoV-19/SouthAfrica/NICD-N10245/2021 | EPI_ISL_2988878 | Gauteng | Unknown | Highlighter plot |
| hCoV-19/SouthAfrica/NICD-N10255/2021 | EPI_ISL_2988405 | Gauteng | Unknown | Tree, temporal analysis, highlighter plot |
| hCoV-19/SouthAfrica/NICD-N10334/2021 | EPI_ISL_3149307 | Gauteng | Unknown | Tree, temporal analysis, highlighter plot |
| hCoV-19/SouthAfrica/NICD-N10596/2021 | EPI_ISL_2988409 | Gauteng | Unknown | Tree, temporal analysis, highlighter plot |
| hCoV-19/SouthAfrica/NICD-N11018/2021 | EPI_ISL_3149312 | Limpopo | Unknown | Highlighter plot |
| hCoV-19/SouthAfrica/NICD-N11037/2021 | EPI_ISL_3149313 | Limpopo | Unknown | Tree, temporal analysis, highlighter plot |
| hCoV-19/SouthAfrica/NICD-N11134/2021 | EPI_ISL_3237084 | Gauteng | Unknown | Tree, temporal analysis, highlighter plot |
| hCoV-19/SouthAfrica/NICD-N11146/2021 | EPI_ISL_3237092 | Gauteng | Unknown | Tree, temporal analysis, highlighter plot |
| hCoV-19/SouthAfrica/NICD-N11155/2021 | EPI_ISL_3237098 | Gauteng | Unknown | Tree, temporal analysis, highlighter plot |
| hCoV-19/SouthAfrica/NICD-N11159/2021 | EPI_ISL_3236951 | Gauteng | Unknown | Tree, temporal analysis, highlighter plot |
| hCoV-19/SouthAfrica/NICD-N11162/2021 | EPI_ISL_3237100 | Gauteng | Unknown | Tree, temporal analysis, highlighter plot |
| hCoV-19/SouthAfrica/NICD-N11163/2021 | EPI_ISL_3236953 | Gauteng | Unknown | Tree, temporal analysis, highlighter plot |
| hCoV-19/SouthAfrica/NICD-N11200/2021 | EPI_ISL_3101505 | Gauteng | Unknown | Tree, temporal analysis, highlighter plot |
| hCoV-19/SouthAfrica/NICD-N11206/2021 | EPI_ISL_3149299 | Gauteng | Unknown | Tree, temporal analysis, highlighter plot |
| hCoV-19/SouthAfrica/NICD-N11223/2021 | EPI_ISL_3149300 | Gauteng | Unknown | Tree, temporal analysis, highlighter plot |
| hCoV-19/SouthAfrica/NICD-N11230/2021 | EPI_ISL_3149301 | Gauteng | Unknown | Tree, temporal analysis, highlighter plot |
| hCoV-19/SouthAfrica/NICD-N11267/2021 | EPI_ISL_3074033 | Gauteng | Unknown | Tree, temporal analysis, highlighter plot |
| hCoV-19/SouthAfrica/NICD-N11301/2021 | EPI_ISL_3149306 | Gauteng | Yes | Tree, temporal analysis, highlighter plot |
| hCoV-19/SouthAfrica/NICD-N11737/2021 | EPI_ISL_3838300 | Limpopo | Unknown | Tree, temporal analysis, highlighter plot |
| hCoV-19/SouthAfrica/NICD-N11765/2021 | EPI_ISL_3838321 | Northern Cape | Unknown | Tree, temporal analysis, highlighter plot |
| hCoV-19/SouthAfrica/NICD-N11891/2021 | EPI_ISL_3411681 | Gauteng | Unknown | Tree, temporal analysis, highlighter plot |
| hCoV-19/SouthAfrica/NICD-N12028/2021 | EPI_ISL_3451144 | Limpopo | Unknown | Tree, temporal analysis, highlighter plot |
| hCoV-19/SouthAfrica/NICD-N12106/2021 | EPI_ISL_3838375 | Eastern Cape | Unknown | Tree, temporal analysis, highlighter plot |
| hCoV-19/SouthAfrica/NICD-N12157/2021 | EPI_ISL_3237233 | Eastern Cape | Unknown | Tree, temporal analysis, highlighter plot |
| hCoV-19/SouthAfrica/NICD-N12240/2021 | EPI_ISL_3411589 | Gauteng | Unknown | Tree, temporal analysis, highlighter plot |
| hCoV-19/SouthAfrica/NICD-N12264/2021 | EPI_ISL_3237237 | Limpopo | Unknown | Tree, temporal analysis, highlighter plot |
| hCoV-19/SouthAfrica/NICD-N12618/2021 | EPI_ISL_3451195 | Gauteng | Unknown | Tree, temporal analysis, highlighter plot |
| hCoV-19/SouthAfrica/NICD-N12640/2021 | EPI_ISL_3451214 | Gauteng | Unknown | Tree, temporal analysis, highlighter plot |
| hCoV-19/SouthAfrica/NICD-N12648/2021 | EPI_ISL_3451222 | Limpopo | Unknown | Tree, temporal analysis, highlighter plot |
| hCoV-19/SouthAfrica/NICD-N12735/2021 | EPI_ISL_3451295 | Northern Cape | Unknown | Tree, temporal analysis, highlighter plot |
| hCoV-19/SouthAfrica/NICD-N12741/2021 | EPI_ISL_3451301 | Northern Cape | Unknown | Tree, temporal analysis, highlighter plot |
| hCoV-19/SouthAfrica/NICD-N12752/2021 | EPI_ISL_3411463 | Gauteng | Unknown | Tree, temporal analysis, highlighter plot |
| hCoV-19/SouthAfrica/NICD-N12787/2021 | EPI_ISL_3411467 | Gauteng | Unknown | Tree, temporal analysis, highlighter plot |
| hCoV-19/SouthAfrica/NICD-N12833/2021 | EPI_ISL_3411457 | Limpopo | Unknown | Tree, temporal analysis, highlighter plot |
| hCoV-19/SouthAfrica/NICD-N13046/2021 | EPI_ISL_3451358 | Limpopo | Unknown | Tree, temporal analysis, highlighter plot |
| hCoV-19/SouthAfrica/NICD-N13052/2021 | EPI_ISL_3451362 | Limpopo | Unknown | Tree, temporal analysis, highlighter plot |
| hCoV-19/SouthAfrica/NICD-N13062/2021 | EPI_ISL_3451369 | Limpopo | Unknown | Tree, temporal analysis, highlighter plot |
| hCoV-19/SouthAfrica/NICD-N13072/2021 | EPI_ISL_3451378 | Limpopo | Unknown | Tree, temporal analysis, highlighter plot |
| hCoV-19/SouthAfrica/NICD-N13090/2021 | EPI_ISL_3451391 | Limpopo | Unknown | Tree, temporal analysis, highlighter plot |
| hCoV-19/SouthAfrica/NICD-N13117/2021 | EPI_ISL_3411459 | Limpopo | Unknown | Tree, temporal analysis, highlighter plot |
| hCoV-19/SouthAfrica/NICD-N13178/2021 | EPI_ISL_3643965 | Mpumalanga | Unknown | Tree, temporal analysis, highlighter plot |
| hCoV-19/SouthAfrica/NICD-N13199/2021 | EPI_ISL_3643862 | Mpumalanga | Unknown | Tree, temporal analysis, highlighter plot |
| hCoV-19/SouthAfrica/NICD-N13207/2021 | EPI_ISL_3643860 | Mpumalanga | Unknown | Tree, temporal analysis, highlighter plot |
| hCoV-19/SouthAfrica/NICD-N13250/2021 | EPI_ISL_3411458 | Limpopo | Unknown | Tree, temporal analysis, highlighter plot |
| hCoV-19/SouthAfrica/NICD-N13314/2021 | EPI_ISL_3643903 | Limpopo | Unknown | Tree, temporal analysis, highlighter plot |
| hCoV-19/SouthAfrica/NICD-N13319/2021 | EPI_ISL_3717995 | Limpopo | Unknown | Tree, temporal analysis, highlighter plot |
| hCoV-19/SouthAfrica/NICD-N13558/2021 | EPI_ISL_4030025 | Northern Cape | Unknown | Tree, temporal analysis, highlighter plot |
| hCoV-19/SouthAfrica/NICD-N13561/2021 | EPI_ISL_4029946 | Northern Cape | Unknown | Tree, temporal analysis, highlighter plot |
| hCoV-19/SouthAfrica/NICD-N13574/2021 | EPI_ISL_4030281 | Northern Cape | Unknown | Highlighter plot |
| hCoV-19/SouthAfrica/NICD-N13584/2021 | EPI_ISL_4030022 | Northern Cape | Unknown | Tree, temporal analysis, highlighter plot |
| hCoV-19/SouthAfrica/NICD-N13587/2021 | EPI_ISL_4029964 | Northern Cape | Unknown | Tree, temporal analysis, highlighter plot |
| hCoV-19/SouthAfrica/NICD-N13588/2021 | EPI_ISL_4029943 | Northern Cape | Unknown | Tree, temporal analysis, highlighter plot |
| hCoV-19/SouthAfrica/NICD-N13589/2021 | EPI_ISL_4029948 | Northern Cape | Unknown | Tree, temporal analysis, highlighter plot |
| hCoV-19/SouthAfrica/NICD-N13597/2021 | EPI_ISL_4029923 | North West | Unknown | Tree, temporal analysis, highlighter plot |
| hCoV-19/SouthAfrica/NICD-N13667/2021 | EPI_ISL_3838489 | Limpopo | Unknown | Tree, temporal analysis, highlighter plot |
| hCoV-19/SouthAfrica/NICD-N13682/2021 | EPI_ISL_3643842 | North West | Unknown | Tree, temporal analysis, highlighter plot |
| hCoV-19/SouthAfrica/NICD-N13716/2021 | EPI_ISL_3838512 | Northern Cape | Unknown | Tree, temporal analysis, highlighter plot |
| hCoV-19/SouthAfrica/NICD-N13719/2021 | EPI_ISL_3838515 | Northern Cape | Unknown | Tree, temporal analysis, highlighter plot |
| hCoV-19/SouthAfrica/NICD-N13725/2021 | EPI_ISL_3838520 | Northern Cape | Unknown | Tree, temporal analysis, highlighter plot |
| hCoV-19/SouthAfrica/NICD-N13748/2021 | EPI_ISL_3838541 | Northern Cape | Unknown | Tree, temporal analysis, highlighter plot |
| hCoV-19/SouthAfrica/NICD-N13763/2021 | EPI_ISL_3838556 | Northern Cape | Unknown | Tree, temporal analysis, highlighter plot |
| hCoV-19/SouthAfrica/NICD-N14080/2021 | EPI_ISL_3838621 | Gauteng | Yes | Tree, temporal analysis, highlighter plot |
| hCoV-19/SouthAfrica/NICD-N14096/2021 | EPI_ISL_3838634 | Gauteng | Yes | Tree, temporal analysis, highlighter plot |
| hCoV-19/SouthAfrica/NICD-N14123/2021 | EPI_ISL_3717982 | Gauteng | Yes | Tree, temporal analysis, highlighter plot |
| hCoV-19/SouthAfrica/NICD-N14134/2021 | EPI_ISL_3717911 | Gauteng | Yes | Tree, temporal analysis, highlighter plot |
| hCoV-19/SouthAfrica/NICD-N14135/2021 | EPI_ISL_3718000 | Gauteng | Unknown | Tree, temporal analysis, highlighter plot |
| hCoV-19/SouthAfrica/NICD-N14142/2021 | EPI_ISL_3717932 | Gauteng | Unknown | Tree, temporal analysis, highlighter plot |
| hCoV-19/SouthAfrica/NICD-N14153/2021 | EPI_ISL_3717972 | Gauteng | Yes | Tree, temporal analysis, highlighter plot |
| hCoV-19/SouthAfrica/NICD-N14154/2021 | EPI_ISL_3717993 | Gauteng | Unknown | Tree, temporal analysis, highlighter plot |
| hCoV-19/SouthAfrica/NICD-N14255/2021 | EPI_ISL_3838569 | Free State | Unknown | Tree, temporal analysis, highlighter plot |
| hCoV-19/SouthAfrica/NICD-N8104/2021 | EPI_ISL_2695610 | Mpumalanga | Unknown | Tree, temporal analysis, highlighter plot |
| hCoV-19/SouthAfrica/NICD-N8127/2021 | EPI_ISL_2695631 | Mpumalanga | Unknown | Highlighter plot |
| hCoV-19/SouthAfrica/NICD-N8831/2021 | EPI_ISL_3342730 | Gauteng | Unknown | Tree, temporal analysis, highlighter plot |
| hCoV-19/SouthAfrica/NICD-N8834/2021 | EPI_ISL_3342731 | Gauteng | Unknown | Tree, temporal analysis, highlighter plot |
| hCoV-19/SouthAfrica/NICD-N8841/2021 | EPI_ISL_3342732 | Gauteng | Unknown | Tree, temporal analysis, highlighter plot |
| hCoV-19/SouthAfrica/NICD-N8844/2021 | EPI_ISL_3342733 | Gauteng | Unknown | Tree, temporal analysis, highlighter plot |
| hCoV-19/SouthAfrica/NICD-N9216/2021 | EPI_ISL_3342734 | Gauteng | Unknown | Tree, temporal analysis, highlighter plot |
| hCoV-19/SouthAfrica/NICD-N9250/2021 | EPI_ISL_3342735 | Gauteng | Unknown | Tree, temporal analysis, highlighter plot |
| hCoV-19/SouthAfrica/NICD-N9382/2021 | EPI_ISL_2828749 | Gauteng | Unknown | Tree, temporal analysis, highlighter plot |
| hCoV-19/SouthAfrica/NICD-N9628/2021 | EPI_ISL_2827937 | Limpopo | Unknown | Tree, temporal analysis, highlighter plot |
| hCoV-19/SouthAfrica/NICD-N9826/2021 | EPI_ISL_2942287 | Gauteng | Unknown | Tree, temporal analysis, highlighter plot |
| hCoV-19/SouthAfrica/NICD-N9882/2021 | EPI_ISL_3643966 | Gauteng | Unknown | Tree, temporal analysis, highlighter plot |
| hCoV-19/SouthAfrica/NICD-R10075/2021 | EPI_ISL_4029912 | North West | Unknown | Tree, temporal analysis, highlighter plot |
| hCoV-19/SouthAfrica/NICD-R10186/2021 | EPI_ISL_4030023 | Gauteng | Unknown | Tree, temporal analysis, highlighter plot |
| hCoV-19/SouthAfrica/NICD-R10233/2021 | EPI_ISL_4029941 | Gauteng | Unknown | Tree, temporal analysis, highlighter plot |
| hCoV-19/SouthAfrica/NICD-R10238/2021 | EPI_ISL_4030024 | Gauteng | Unknown | Tree, temporal analysis, highlighter plot |
| hCoV-19/SouthAfrica/NICD-R10269/2021 | EPI_ISL_4030021 | Mpumalanga | Unknown | Tree, temporal analysis, highlighter plot |
| hCoV-19/SouthAfrica/NICD-R10630/2021 | EPI_ISL_3219805 | Gauteng | Unknown | Tree, temporal analysis, highlighter plot |
| hCoV-19/SouthAfrica/NICD-R10925/2021 | EPI_ISL_3219868 | Limpopo | Unknown | Tree, temporal analysis, highlighter plot |
| hCoV-19/SouthAfrica/NICD-R11322/2021 | EPI_ISL_3451544 | KwaZulu-Natal | Unknown | Tree, temporal analysis, highlighter plot |
| hCoV-19/SouthAfrica/NICD-R11374/2021 | EPI_ISL_3451569 | North West | Unknown | Tree, temporal analysis, highlighter plot |
| hCoV-19/SouthAfrica/NICD-R12465/2021 | EPI_ISL_4029935 | Cape Town | Unknown | Tree, temporal analysis, highlighter plot |
| hCoV-19/SouthAfrica/Tygerberg_1419/2021 | EPI_ISL_3118719 | Western Cape | Unknown | Tree, temporal analysis, highlighter plot |
| hCoV-19/SouthAfrica/Tygerberg_1521/2021 | EPI_ISL_3482519 | Western Cape | Unknown | Tree, temporal analysis, highlighter plot |
| hCoV-19/SouthAfrica/UFS-VIRO-NGS-ONP180821NB19/2021 | EPI_ISL_3746811 | Free State | Unknown | Tree, temporal analysis, highlighter plot |
| hCoV-19/SouthAfrica/UFS-VIRO-NGS-ONP180821NB28/2021 | EPI_ISL_3746772 | Free State | Unknown | Tree, temporal analysis, highlighter plot |
| hCoV-19/SouthAfrica/UFS-VIRO-NGS-ONP180821NB45/2021 | EPI_ISL_3746842 | Free State | Unknown | Tree, temporal analysis, highlighter plot |
| hCoV-19/SouthAfrica/UFS-VIRO-NGS-ONP180821NB47/2021 | EPI_ISL_3746871 | Free State | Unknown | Highlighter plot |
| hCoV-19/SouthAfrica/UFS-VIRO-NGS-ONP180821NB50/2021 | EPI_ISL_3746752 | Free State | Unknown | Tree, temporal analysis, highlighter plot |
| hCoV-19/SouthAfrica/UFS-VIRO-NGS-ONP180821NB53/2021 | EPI_ISL_3746788 | Free State | Unknown | Tree, temporal analysis, highlighter plot |
| hCoV-19/SouthAfrica/UFS-VIRO-NGS-ONP180821NB71/2021 | EPI_ISL_3746874 | Free State | Unknown | Highlighter plot |
| hCoV-19/SouthAfrica/VIDA-KRISP-K018954/2021 | EPI_ISL_2841668 | Gauteng | Unknown | Tree, temporal analysis, highlighter plot |
| hCoV-19/SouthAfrica/VIDA-KRISP-K018963/2021 | EPI_ISL_2841677 | Gauteng | Unknown | Tree, temporal analysis, highlighter plot |
| hCoV-19/SouthAfrica/VIDA-KRISP-K020434/2021 | EPI_ISL_3729072 | Gauteng | Yes | Tree, temporal analysis, highlighter plot |
| hCoV-19/SouthAfrica/VIDA-KRISP-K020445/2021 | EPI_ISL_3729227 | Gauteng | Yes | Tree, temporal analysis, highlighter plot |

**Supplementary Table 2:**
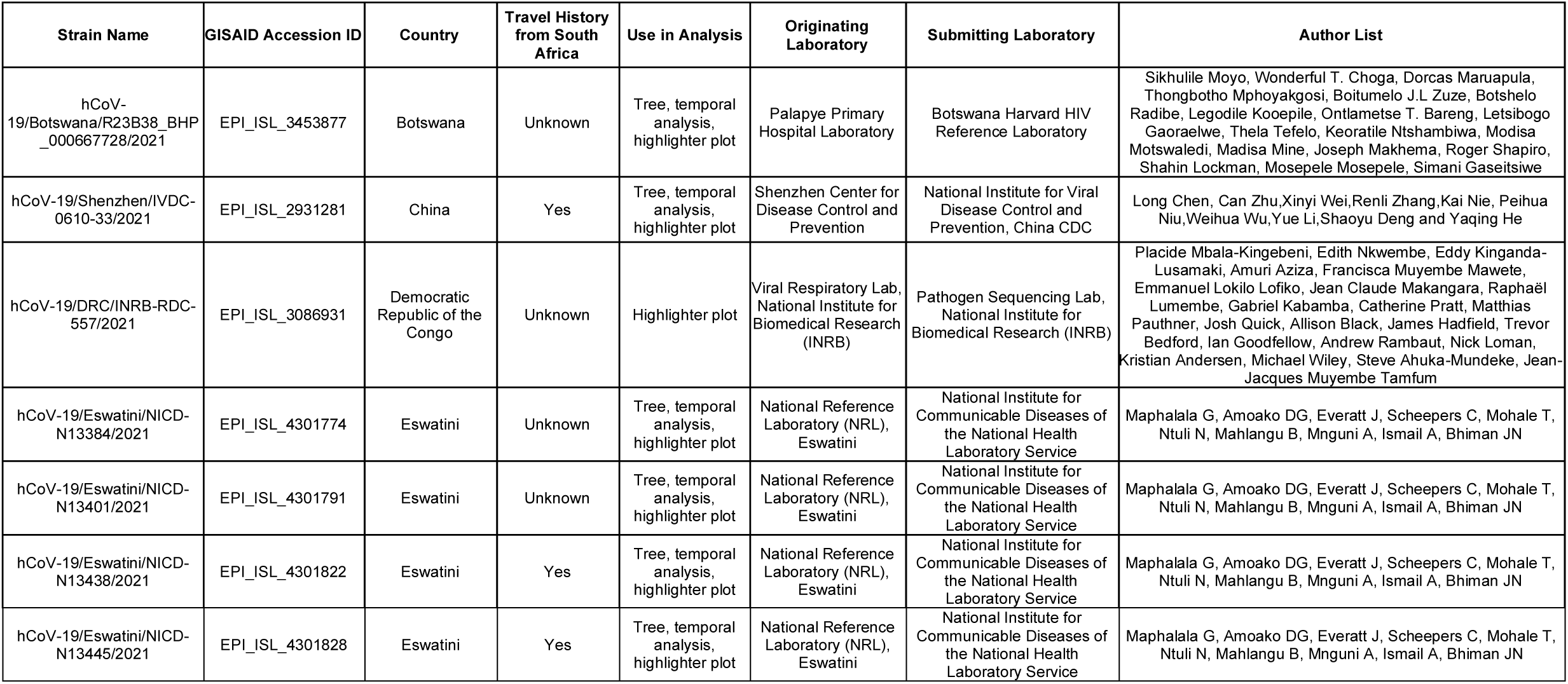
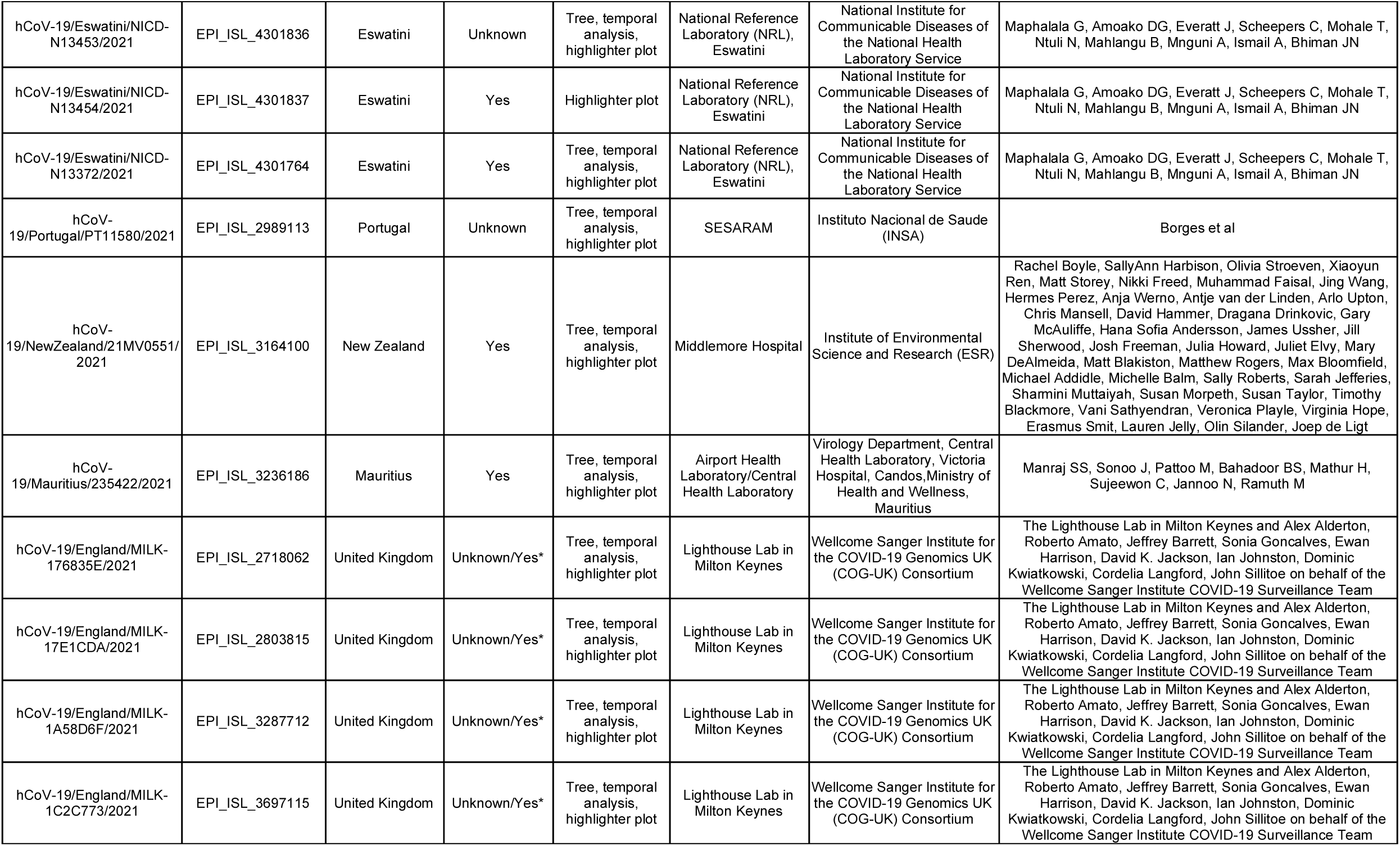
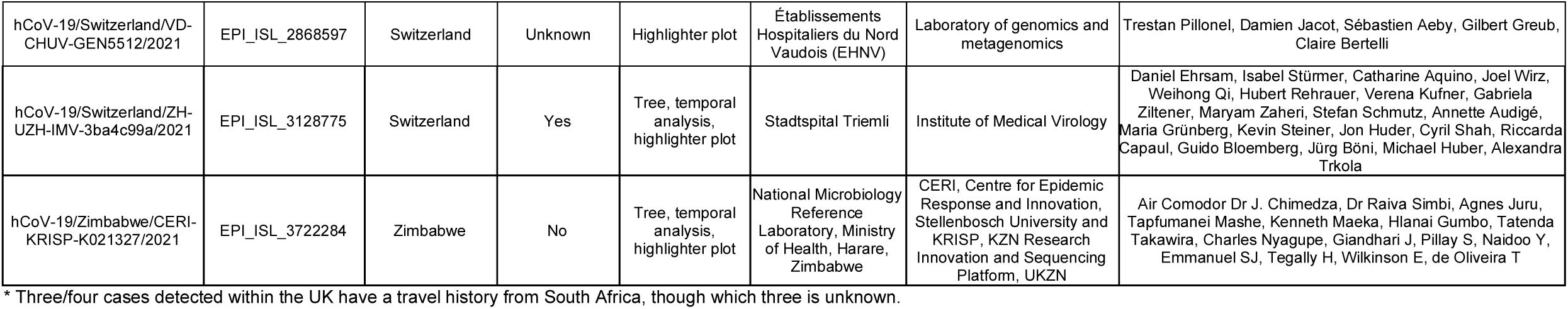
Reference set of C.1.2 genomes on GISAID from other countries. We gratefully acknowledge the following authors from the originating laboratories responsible for obtaining the specimen, as well as the submitting laboratories where the genomes were generated and shared via GISAID, on which this research is based. All submitters of data may be contacted via www.gisaid.org. Provided are the GISAID strain name and GISAID_EPI_ISL accession numbers, authors (listed according to how they were provided on GISAID), travel status and use in various analyses for non-South African C.1.2 samples deposited into GISAID as of September, 10, 2021.

